# Comparative Analysis of Health Care Use and Costs for Orthobiologic versus Surgical Treatments in Economically High-Impact Knee Conditions

**DOI:** 10.64898/2026.02.27.26347270

**Authors:** Trevor A. Lentz, Joshua Burrows, Amanda Brucker, A. Ian Wong, Laura Qualls, Rekha Divakaran, Chris Centeno, Tim Suther, Laine Thomas

## Abstract

**Background:** Total knee arthroplasty (TKA), partial knee arthroplasty (pKA), and arthroscopic meniscectomy are among the most commonly performed procedures for knee osteoarthritis and degenerative meniscal tears in the United States, yet concerns persist regarding overuse, variable clinical benefit, and high costs. Orthobiologic treatments, including platelet-rich plasma (PRP) and bone marrow aspirate concentrate (BMAC), have emerged as less invasive alternatives, but downstream health care resource use (HCRU) and costs associated with these treatments relative to surgery are not well established.

**Methods:** We conducted a retrospective, observational cohort study using linked commercial insurance claims data and a national orthobiologic treatment registry to compare downstream HCRU and costs following orthobiologic versus surgical treatment of knee conditions. Two comparisons were evaluated separately: (1) PRP versus arthroscopic meniscectomy among patients with degenerative meniscal pathology and minimal osteoarthritis, and (2) BMAC with or without PRP versus TKA or pKA among patients with knee osteoarthritis. Eligible procedures occurred between 2016 and 2023. Propensity score matching was used to balance demographic and clinical confounders. Primary outcomes were total health care costs at 12 and 24 months post-procedure, with exploratory analyses at 36 and 48 months. Costs were estimated using multiple approaches, including Medicare-based estimates, commercial payer estimates, and aggregate allowed amounts. HCRU outcomes included outpatient visits, physical therapy, imaging, opioid use, repeat injections, and subsequent surgery.

**Results:** After matching, analyses included 167 PRP-treated patients matched to 1,670 meniscectomy patients and 165 BMAC/PRP-treated patients matched to 1,650 TKA/pKA patients, with good balance across pre-specified confounders. Progression to subsequent surgery after orthobiologic treatment was rare at 12 and 24 months in both cohorts. Compared with TKA/pKA, BMAC/PRP was associated with lower overall health care use for several services, including outpatient visits, physical therapy, knee radiographs, and opioid prescriptions, although magnetic resonance imaging was more frequent following orthobiologic treatment. Total costs within 12 and 24 months were consistently higher for TKA/pKA than for BMAC/PRP across all costing methods. In the PRP versus meniscectomy comparison, differences in health care use were modest, and costs were similar or lower for PRP depending on the costing approach. Exploratory analyses through 48 months showed similar patterns, with persistently low rates of subsequent surgery after orthobiologic treatment and generally higher cumulative costs following surgical intervention.

**Conclusions:** In this real-world, propensity-matched analysis of commercially insured patients, orthobiologic treatments with PRP or BMAC were associated with similar or lower downstream health care costs compared with commonly performed surgical alternatives for selected patients with degenerative meniscal tears or knee osteoarthritis. Progression to surgery following orthobiologic treatment was uncommon through two years and remained low in longer-term exploratory analyses. These findings support the consideration of orthobiologic therapies as potentially lower-cost alternatives to surgery for appropriately selected patients and may inform shared decision-making and payer policy.

## BACKGROUND

Total knee arthroplasty (TKA) and arthroscopic meniscectomy are among the most common surgical procedures for knee pathology in the United States.^4,27,36,38,40^ Although both procedures remain widely accepted for managing advanced osteoarthritis and meniscal tears, respectively, their use has come under increasing scrutiny.^1,21,34,35^ Concerns are focused on the appropriateness and overuse of these interventions, particularly when less invasive options have not been fully exhausted or where clinical benefit may be limited. Research indicates that up to one-third of total knee arthroplasty (TKA) procedures may be unnecessary^34^, and nearly 25% of patients report bothersome or high-impact chronic pain following surgery^14^. For people with degenerative meniscal tears, meniscectomy has been shown to be no more beneficial than sham surgery^37^ or exercise^3,30^. These findings have prompted calls for a more judicious approach to surgical intervention for many degenerative joint conditions, particularly as efforts intensify to reduce musculoskeletal care costs.

Surgical procedures such as TKA and meniscectomy carry inherent risks, including infection, thromboembolic events, and prolonged recovery, and impose substantial financial burdens on patients and payers.^17,39,41^ As a result, there is increasing interest in less costly, nonsurgical alternatives that may offer symptom relief with reduced risk. Orthobiologic therapies are one such option. Two types of orthobiologics, platelet-rich plasma (PRP) and bone marrow aspirate concentrate (BMAC), have demonstrated strong potential for the safe and effective treatment of various musculoskeletal conditions.^2,7,10,18,19,29^ Platelets and cells concentrated in PRP and BMAC release cytokines, exosomes, and growth factors that are believed to initiate intercellular signaling and promote tissue repair.^9,23^ BMAC contains mesenchymal stem cells (MSCs), hematopoietic cells, and anti-inflammatory cytokines which support skeletal tissue healing.^16^ As multipotent cells, MSCs have the capacity to differentiate into cartilage and bone, contributing to cartilage regeneration and subchondral bone remodeling.^5,22,43^

Although PRP and BMAC are most commonly used to treat cartilage-related and meniscal conditions, their overall use in managing musculoskeletal conditions remains low (5.9 to 7.9 per 1000 patients).^42^ Preliminary evidence suggests these therapies can improve pain and function in patients with degenerative meniscal pathology and osteoarthritis, though their long-term impact on disease progression and health care use remains unclear.^2,10–12^ Orthobiologic treatments have been proposed as alternatives to surgery for some conditions; however, few studies have directly compared orthobiologic therapies with surgical intervention, with most comparisons instead limited to other non⍰surgical treatments.^8,15,20,25,32^

The primary value proposition of orthobiologic treatment is its potential to offer a less costly and lower risk alternative to surgery for degenerative joint conditions, but this hypothesis has not been rigorously tested. Although orthobiologic treatment may offer an effective alternative to surgery for some conditions, it may also increase downstream costs related to repeat injections, additional imaging, or eventual surgical intervention, raising concerns about overall economic feasibility. As payer policies evolve and musculoskeletal care costs continue to rise, understanding how orthobiologics perform from a cost and health care use standpoint relative to surgery is essential for informed decision-making.

This study compares downstream health care use and costs among patients with knee osteoarthritis and/or degenerative meniscal tears that meet diagnostic criteria for orthopedic surgery or who are otherwise functionally disabled and elect to undergo either BMAC/PRP injection or surgical intervention (arthroplasty or meniscectomy, respectively). The primary endpoint is aggregate cost over 12 and 24 months with exploratory analyses at 36 and 48 months post procedure. By analyzing real-world data, we aim to understand if PRP and BMAC procedures serve as a less costly alternative to surgery or contribute to increased health care burden. These insights are critical for guiding clinical decision-making, shaping payer coverage policies, and reducing low-value care in musculoskeletal health.

## METHODS

### Study design and population

This is a retrospective, observational study using health care billing and claims data to compare cost and resource utilization outcomes after knee BMAC or PRP injection or knee surgery. Two study populations were evaluated separately: those undergoing 1) autologous BMAC injection with or without PRP (BMAC/PRP) versus TKA or partial knee arthroplasty (pKA) for management of knee osteoarthritis; and 2) PRP injection versus meniscectomy in patients eligible for both. All procedures occurred between 2016 and 2023. All patients were enrolled in a commercial health insurance plan.

### Data sources

Health care use including facility, profession and pharmacy claims and financial fields for all associated claims data was provided by a nationwide commercial insurance provider, through their research data platform. Information on BMAC and PRP procedures was extracted from the Regenexx patient registry and included procedure date, joint treated, location treated, and type of injection. Other than these specific procedural variables, all data including covariates and outcomes were obtained consistently from the commercial claims data source for all comparator arms. The procedural information from Regenexx was linked to the commercial claims database using a privacy-preserving, token-based method performed by Datavant, Inc. No identifiable information, except dates of health care services, was shared across organizations, and the combined analytic dataset was de-identified by expert determination. Analyses were conducted within a secure cloud-based analytic enclave that prevents data exfiltration; only privacy-screened summary results are permitted to leave the environment. This study was approved by the Duke University IRB Pro00116431.

### Eligibility criteria

Procedures were eligible if 1) they received an eligible orthobiologic treatment of the knee (BMAC and/or PRP; Regenexx, Des Moines, IA) or underwent unilateral surgery (TKA, pKA, meniscectomy); 2) procedure occurs between 2016 and 2023; 3) patient was age 18 years or older at the time of the procedure; 4) the patient had commercial coverage from 12 months prior to procedure to at least 12 months after procedure; 5) patient had at least one outpatient encounter (claim) in the 12 months prior to the procedure and at least one outpatient encounter (claim) in the 12 months after the procedure; and 6) procedure was elective and preceded by a relevant clinical diagnosis (Appendix A). Surgical CPT codes were used to identify procedures (Appendix B). Complicated cases involving multiple, distinct types of surgery during the same encounter were not included (e.g. quadriceps repair and TKA)

Exclusion criteria were: 1) patients who had multiple birthdates or sexes in the commercial payer data; 2) patients with ineligible diagnoses (Appendix C), e.g., inflammatory syndrome diagnosis, infectious arthropathies, or bone cancer in the 12 months prior to the procedure; 3) previous knee surgery in the 12 months prior to the procedure; or 4) evidence of receiving a Monoclonal Antibody (MAb) drug in the three months prior to the procedure. We excluded patients with evidence of corticosteroids (oral or injection) within 2 months prior to a BMAC/PRP or PRP procedure, as this is contra-indicated for either BMAC/PRP or PRP. If there were multiple eligible procedures for a patient, we took the first that met the eligibility criteria.

### Cohorts and comparators

The comparators of interest were: (1) TKA/pKA versus BMAC with or without PRP (BMAC/PRP) in patients who would have been good candidates for both TKA/pKA and BMAC/PRP; and (2) Meniscectomy versus PRP in patients who would have been a good candidate for both meniscectomy and PRP. Details of BMAC/PRP and PRP treatments, including their preparation and administration, are provided elsewhere.^6,13^ In practice, whether a patient is a good candidate for these procedures is based on objective criteria as well as subjective judgment. Specifically, each orthobiologic procedure (BMAC/PRP or PRP) predominantly corresponds to an alternative surgical procedure that could have theoretically been chosen instead. PRP injections are most commonly used for treatment of meniscal tears (meniscectomy comparator), while autologous BMAC/PRP is typically used for treatment of osteoarthritis (TKA/pKA comparator).

Based on this contextual information, we used two strategies to define comparable cohorts and minimize confounding. First, we applied relevant inclusion/exclusion criteria. To be included in the comparison of TKA/pKA versus BMAC/PRP, patients had to meet study eligibility criteria and have a knee osteoarthritis diagnosis (Appendix B) in the 12 months prior to procedure. This aligns with the expectation that osteoarthritis plus BMAC, rather than PRP alone, are minimally necessary for potential comparability to TKA/PKA. To be included in the meniscectomy surgery versus PRP cohort, patients had to meet study eligibility criteria and have no evidence of major osteoarthritis defined as ≤ 1 visit for osteoarthritis in the 12 months prior to procedure, and no evidence of anterior cruciate or medial collateral ligament (ACL/MCL) injury (Appendix B). These criteria are meant to exclude reasons for PRP that would not be treated by meniscectomy. Second, we used propensity score matching to refine comparability (as described below).

### Confounders

We derived and reported as many potential confounders as possible. Demographic information included age, sex, and geographic region. Health related characteristics included injury to the treated knee, orthopedic diagnoses, other musculoskeletal conditions not exclusive to the treated knee, comorbidities, pain medication prescriptions, intervention history, health seeking behavior, and other severity proxies (e.g., number of outpatient encounters due to osteoarthritis OA). For derived variables that involve diagnosis or procedures codes, we counted any occurrence in the 12-month period prior to the date of the index procedure (excluding the date of the index procedure itself). Confounders were pre-specified prior to conducting any analysis of outcome data. Specifically, we identified variables that were different between the treatment groups and had a plausible impact on post-procedural cost outcomes (listed in Appendix D). Variables that were rare, already well-balanced, or highly correlated with an included variable were not used within the propensity score matching process (described below). Diagnostic procedures, like X-ray an MRI, were part of preoperative work up for surgery and diagnoses required for surgical reimbursement (meniscal for meniscectomy) were highly correlated with surgery. These variables were bundled with the choice of surgery (reverse causation) and were not treated as confounders. Descriptive data are reported on all variables regardless of whether they were regarded as confounders or used for matching.

### Health care resource use (HCRU)

We measured HCRU by interactions with the healthcare system, i.e. claims for healthcare encounters in the large commercial data. HCRU was captured longitudinally from the day after the procedure end date until 12, 24, 36, and 48 months after the procedure start date. Encounter types included: outpatient (non-physical therapy) visits, outpatient physical therapy, radiology, surgical procedures (on the same knee), subsequent BMAC or PRP procedures, emergency or inpatient care, and opioid medications. When the knee side was unspecified, we counted it as the same knee for all procedure types. Details of HCRU attribution and episode definitions are in Appendix E.

### Cost outcomes

The primary outcome was the aggregate costs associated with all relevant HCRU services (defined above) measured at 12 and 24 months post-procedure (details in Appendix F). Due to low sample size of patients with extended commercial payer coverage, we assessed cost outcomes out to 36 and 48 months post-procedure as an exploratory aim. We obtained cost estimates under two payment models, Medicare and commercial payer. External Medicare costs were estimated using values from the literature (See Appendix F for a table of costs and associated references). To estimate commercial payer costs, we applied three methods. First, Medicare-based costs were multiplied by 2, which is an approximate conversion of Medicare to commercial payer costs based on an average of multipliers across different cost settings.^26^ Second, we used the available commercial payer cost data and computed average allowed amounts per episode of care (day for outpatient services, and full episode of care for inpatient/emergency). Each HCRU was assigned a cost value, based on this average (Appendix F for table of assigned costs). Finally, we used a direct method of summing the actual costs (allowed amounts) of relevant HCRU services for those matched patients who were included in the analysis. This last approach introduced some outlying costs that appeared likely to be data errors. We therefore added a truncated version of this approach by truncating the costs at the 5^th^ and 95^th^ percentile prior to making comparisons.

### Statistical analysis

Descriptive data are reported overall and separately for the TKA/pKA surgical cohort and meniscectomy surgical cohort, including frequency counts and percentages for categorical variables and mean, standard deviation, median, interquartile range, minimum and maximum for continuous variables. We developed propensity score models and propensity scores for the purpose of creating comparability between those receiving BMAC/PRP versus surgical procedures. We developed two separate propensity models for the two treatment comparisons: BMAC/PRP versus TKA/pKA and PRP versus meniscectomy. All comparative analysis were conducted separately for the TKA/pKA and meniscectomy surgical cohorts.

To obtain propensity scores, we fit a logistic regression model for all individuals in the comparison set with a binary outcome of receiving surgical or orthobiologic treatment. For each patient, the linear predictor (X*β) was estimated by applying the estimated coefficients from the propensity score model to their individual covariates. We conducted 1:10 greedy matching with respect to the linear predictor, beginning with each patient receiving orthobiologic treatment and seeking 10 matching surgical patients within a caliper of 0.20 times the standard deviation of X*β. Continuous covariates were assessed for non-linearity using restricted cubic splines if any imbalance suggested model mis-specification.

To evaluate the adequacy of the treatment selection model, baseline characteristics of each group were compared following matching. For each pre-specified confounder, absolute standardized mean differences (ASMDs) were calculated. ASMDs below 0.10 indicate good balance and below 0.20 moderate balance.

At each timepoint we described HCRU outcomes between treatment comparator groups in the propensity matched population. P-values are included for the comparison of HCRU outcomes, but our purpose was primarily descriptive and not hypothesis driven. Finally, we compared aggregate costs at the co-primary (12 and 24 months) and exploratory (36 and 48 months) endpoints between treatment comparator groups in the propensity matched population. The distribution of cost across patients was reported by comparator group and the difference in mean costs computed along with a 95% confidence interval for the difference.

Due to requirements for privacy protection, cell counts less than 11 were suppressed and are reported as <11. This compromised our ability to report and compare rates of rare outcomes. Therefore, in an exploratory analysis, we relaxed inclusion criteria to remove the requirement for continuous coverage, at least one outpatient claim, and a relevant orthopedic diagnosis in the 12 months prior to the procedure (details in Appendix P). This doubled the sample size of Regenexx patients and allowed specific counts of some rare events to be unmasked. However, without detailed medical history it was not possible to establish comparability between these patients and a particular surgery.

All analyses were conducted in R (version 4.4.2). Significance level of alpha = 0.05 was used for all comparisons and confidence intervals. No multiple testing adjustments were performed.

## RESULTS

After applying eligibility criteria and prior to matching, we identified 167 patients who received PRP and met criteria for comparability to meniscectomy, along with 116,473 patients who underwent meniscectomy, and 185 patients who received BMAC/PRP injection and met criteria for comparability to TKA/pKA, along with 97,327 patients who underwent TKA/pKA. (Table 1, Appendix H & I). After matching, the sample included 167 patients who received PRP injection matched to 1670 patients who underwent meniscectomy and 165 patients who received BMAC/PRP matched to 1650 patients who underwent TKA/pKA. All PRP patients were successfully matched with 10 meniscectomy patients, whereas 20 BMAC/PRP patients did not have comparable matches among TKA/pKA and were excluded by the matching process.

**Table 1.** Cohort flowchart.

| **Inclusion/exclusion criteria** | **Orthobiologic Procedures** | **TKA/pKA Procedures** | **Meniscectomy Procedures** |
| --- | --- | --- | --- |
| Starting population: Receiving eligible procedures | 3833 | 340226 | 452957 |
| Procedure occurred between 1/1/2016 and 12/31/2023 | 3553 | 323702 | 438312 |
| Exclude patients who have multiple DOBs or sexes in commercial claims data | 3553 | 323691 | 438301 |
| Patients age 18 or older at the time of the procedure | 3503 | 323649 | 425899 |
| 12 months of continuous enrollment with commercial insurer both before and after the procedure | 905 | 173995 | 229540 |
| At least one outpatient claim in the 12 months prior to the procedure | 878 | 173989 | 229537 |
| At least one outpatient claim in the 12 months after the procedure | 863 | 173976 | 229537 |
| **Procedure is elective and preceded by a relevant clinical diagnosis** |  |  |  |
| At least one relevant diagnosis in an outpatient setting in the 12 months prior to the procedure | 756 | 173936 | 229526 |
| If ER care in the 3 months prior to the procedure, at least one relevant diagnosis in an outpatient setting after the ER care but before the procedure | 748 | 171875 | 227736 |
| If occurs in hospital, occurs on first day of service | 748 | 169449 | 227687 |
| **Exclude patients with clinical contra-indications** |  |  |  |
| Patients with inflammatory syndrome diagnosis in the 12 months prior to the procedure | 690 | 143851 | 205860 |
| Ineligible connective tissue disorder diagnosis in the 12 months prior to the procedure | 679 | 141451 | 202205 |
| Ineligible joint disorder diagnosis in the 12 months prior to the procedure | 679 | 141165 | 201806 |
| Ineligible injuries in the 12 months prior to the procedure | 675 | 140236 | 200822 |
| Infectious arthropathies in the 12 months prior to the procedure | 673 | 139806 | 200522 |
| Bone cancer in the 12 months prior to the procedure | 672 | 139745 | 200487 |
| **Exclude patients with prior treatment contra-indications** |  |  |  |
| Regenexx patients who had corticosteroids in the 2 months prior to the procedure | 610 | 139745 | 200487 |
| Previous knee surgery in the 12 months prior to the procedure | 589 | 109600 | 183770 |
| Mab medication in the 3 months prior to the procedure | 583 | 108230 | 181865 |
| Surgery patients who had Regenexx at any point | 583 | 108158 | 181760 |
| If a patient had multiple procedures, take the first one | 509 | 98638 | 175487 |
| Comparable patients based on surgery avoided criteria | 352 | 97327 | 116473 |

In both comparison cohorts, pre-specified confounders were well-balanced by matching (ASMDs <0.10) as are nearly all of the measured variables (ASMD<0.20) in Table 2 and 3 except for variables that were either rare or expected to be bundled with surgery (required for pre-op or for reimbursement) and therefore excluded from the set of confounders a-priori.

**Table 2.**
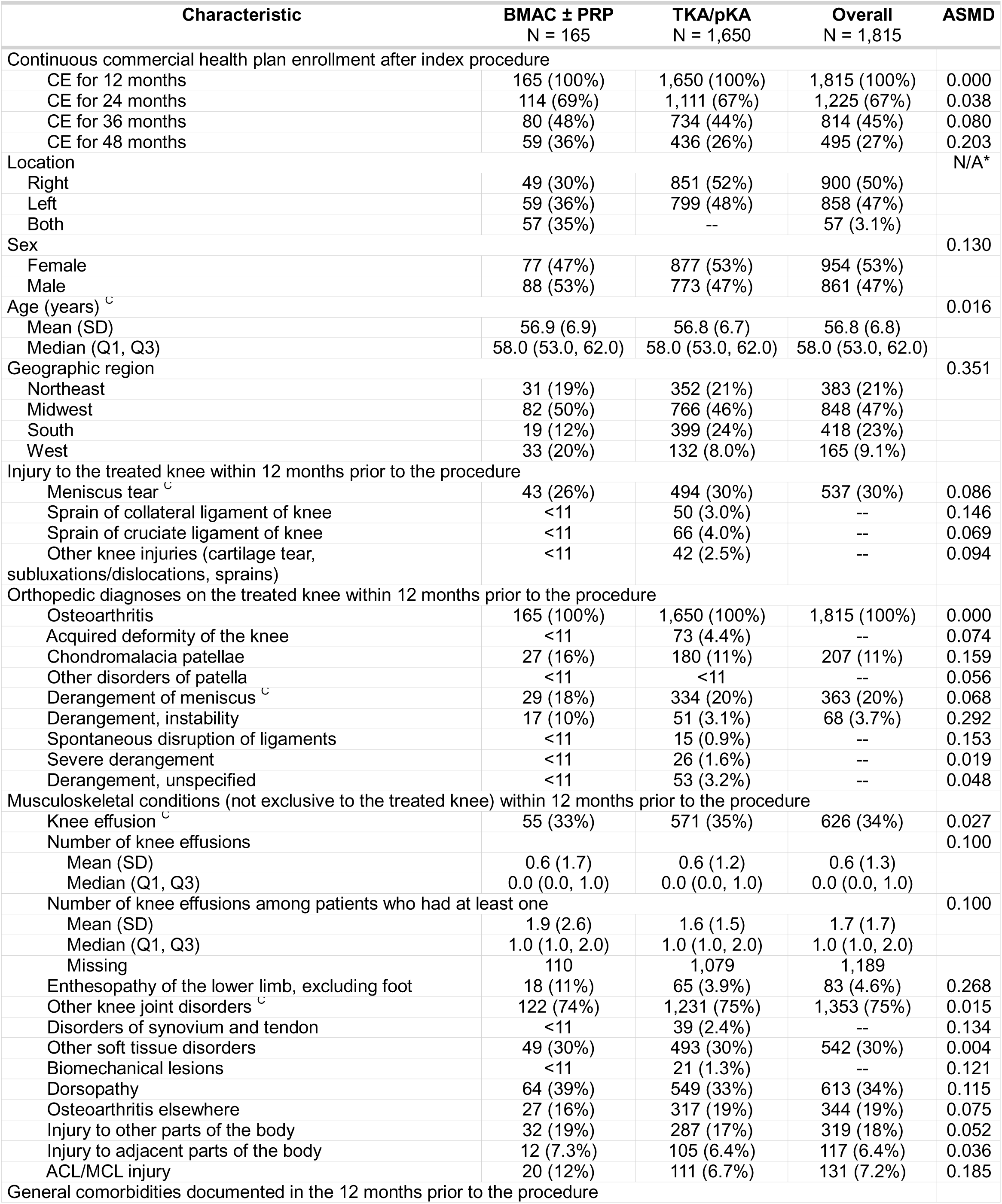
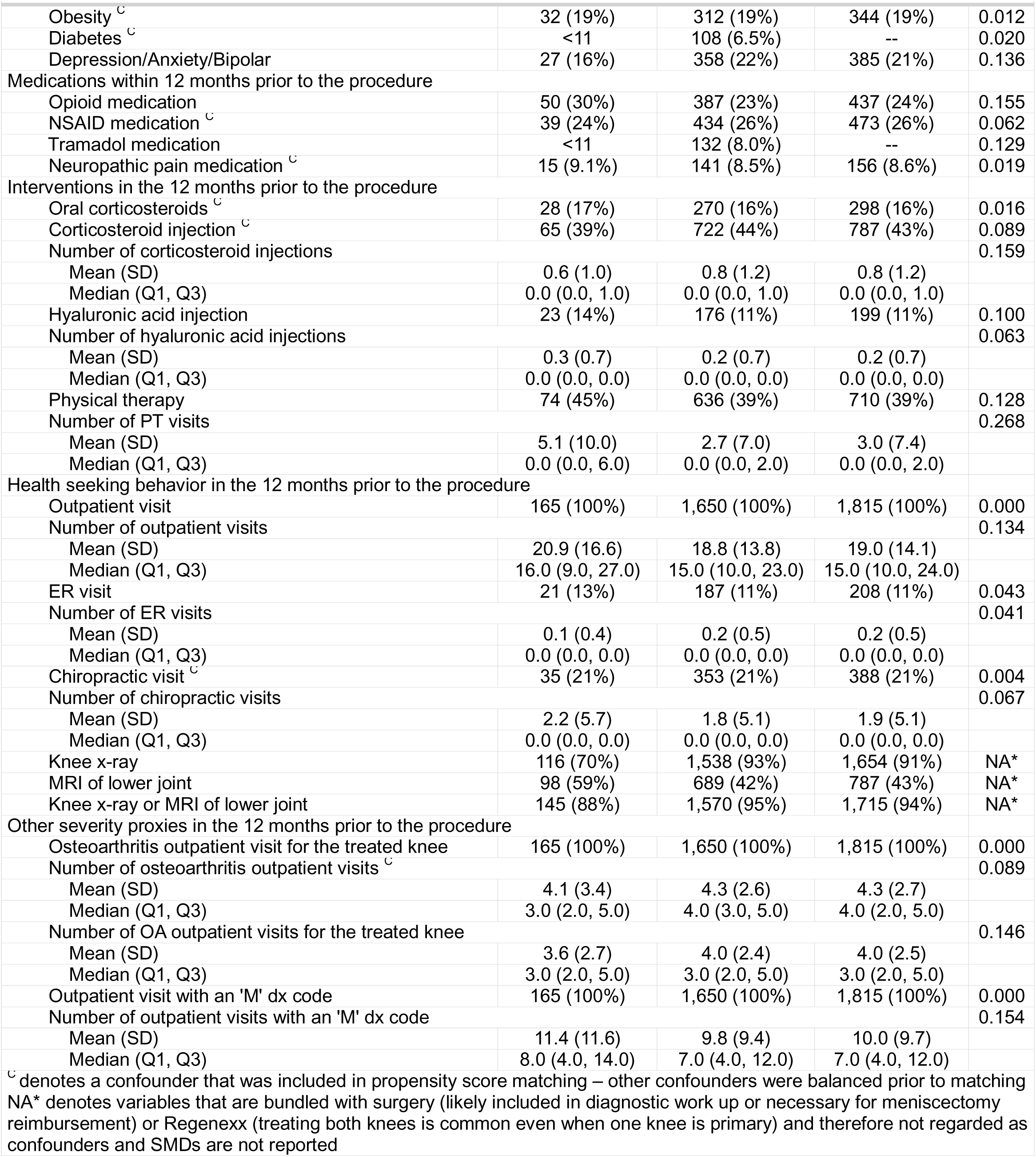
Post-matching demographic and clinical characteristics – BMAC with or without PRP versus TKA/pKA.

| Characteristic | BMAC ± PRP<br>N = 165 | TKA/pKA<br>N = 1,650 | Overall<br>N = 1,815 | ASMD |
| --- | --- | --- | --- | --- |
| Continuous commercial health plan enrollment after index procedure |  |  |  |  |
| CE for 12 months | 165 (100%) | 1,650 (100%) | 1,815 (100%) | 0.000 |
| CE for 24 months | 114 (69%) | 1,111 (67%) | 1,225 (67%) | 0.038 |
| CE for 36 months | 80 (48%) | 734 (44%) | 814 (45%) | 0.080 |
| CE for 48 months | 59 (36%) | 436 (26%) | 495 (27%) | 0.203 |
| Location |  |  |  | N/A* |
| Right | 49 (30%) | 851 (52%) | 900 (50%) |  |
| Left | 59 (36%) | 799 (48%) | 858 (47%) |  |
| Both | 57 (35%) | -- | 57 (3.1%) |  |
| Sex |  |  |  | 0.130 |
| Female | 77 (47%) | 877 (53%) | 954 (53%) |  |
| Male | 88 (53%) | 773 (47%) | 861 (47%) |  |
| Age (years) <sup>∘</sup> |  |  |  | 0.016 |
| Mean (SD) | 56.9 (6.9) | 56.8 (6.7) | 56.8 (6.8) |  |
| Median (Q1, Q3) | 58.0 (53.0, 62.0) | 58.0 (53.0, 62.0) | 58.0 (53.0, 62.0) |  |
| Geographic region |  |  |  | 0.351 |
| Northeast | 31 (19%) | 352 (21%) | 383 (21%) |  |
| Midwest | 82 (50%) | 766 (46%) | 848 (47%) |  |
| South | 19 (12%) | 399 (24%) | 418 (23%) |  |
| West | 33 (20%) | 132 (8.0%) | 165 (9.1%) |  |
| Injury to the treated knee within 12 months prior to the procedure |  |  |  |  |
| Meniscus tear <sup>∘</sup> | 43 (26%) | 494 (30%) | 537 (30%) | 0.086 |
| Sprain of collateral ligament of knee | <11 | 50 (3.0%) | -- | 0.146 |
| Sprain of cruciate ligament of knee | <11 | 66 (4.0%) | -- | 0.069 |
| Other knee injuries (cartilage tear, subluxations/dislocations, sprains) | <11 | 42 (2.5%) | -- | 0.094 |
| Orthopedic diagnoses on the treated knee within 12 months prior to the procedure |  |  |  |  |
| Osteoarthritis | 165 (100%) | 1,650 (100%) | 1,815 (100%) | 0.000 |
| Acquired deformity of the knee | <11 | 73 (4.4%) | -- | 0.074 |
| Chondromalacia patellae | 27 (16%) | 180 (11%) | 207 (11%) | 0.159 |
| Other disorders of patella | <11 | <11 | -- | 0.056 |
| Derangement of meniscus <sup>∘</sup> | 29 (18%) | 334 (20%) | 363 (20%) | 0.068 |
| Derangement, instability | 17 (10%) | 51 (3.1%) | 68 (3.7%) | 0.292 |
| Spontaneous disruption of ligaments | <11 | 15 (0.9%) | -- | 0.153 |
| Severe derangement | <11 | 26 (1.6%) | -- | 0.019 |
| Derangement, unspecified | <11 | 53 (3.2%) | -- | 0.048 |
| Musculoskeletal conditions (not exclusive to the treated knee) within 12 months prior to the procedure |  |  |  |  |
| Knee effusion <sup>∘</sup> | 55 (33%) | 571 (35%) | 626 (34%) | 0.027 |
| Number of knee effusions |  |  |  | 0.100 |
| Mean (SD) | 0.6 (1.7) | 0.6 (1.2) | 0.6 (1.3) |  |
| Median (Q1, Q3) | 0.0 (0.0, 1.0) | 0.0 (0.0, 1.0) | 0.0 (0.0, 1.0) |  |
| Number of knee effusions among patients who had at least one |  |  |  | 0.100 |
| Mean (SD) | 1.9 (2.6) | 1.6 (1.5) | 1.7 (1.7) |  |
| Median (Q1, Q3) | 1.0 (1.0, 2.0) | 1.0 (1.0, 2.0) | 1.0 (1.0, 2.0) |  |
| Missing | 110 | 1,079 | 1,189 |  |
| Enthesopathy of the lower limb, excluding foot | 18 (11%) | 65 (3.9%) | 83 (4.6%) | 0.268 |
| Other knee joint disorders <sup>∘</sup> | 122 (74%) | 1,231 (75%) | 1,353 (75%) | 0.015 |
| Disorders of synovium and tendon | <11 | 39 (2.4%) | -- | 0.134 |
| Other soft tissue disorders | 49 (30%) | 493 (30%) | 542 (30%) | 0.004 |
| Biomechanical lesions | <11 | 21 (1.3%) | -- | 0.121 |
| Dorsopathy | 64 (39%) | 549 (33%) | 613 (34%) | 0.115 |
| Osteoarthritis elsewhere | 27 (16%) | 317 (19%) | 344 (19%) | 0.075 |
| Injury to other parts of the body | 32 (19%) | 287 (17%) | 319 (18%) | 0.052 |
| Injury to adjacent parts of the body | 12 (7.3%) | 105 (6.4%) | 117 (6.4%) | 0.036 |
| ACL/MCL injury | 20 (12%) | 111 (6.7%) | 131 (7.2%) | 0.185 |
| General comorbidities documented in the 12 months prior to the procedure |  |  |  |  |
| Obesity <sup>∘</sup> | 32 (19%) | 312 (19%) | 344 (19%) | 0.012 |
| Diabetes <sup>∘</sup> | <11 | 108 (6.5%) | -- | 0.020 |
| Depression/Anxiety/Bipolar | 27 (16%) | 358 (22%) | 385 (21%) | 0.136 |
| Medications within 12 months prior to the procedure |  |  |  |  |
| Opioid medication | 50 (30%) | 387 (23%) | 437 (24%) | 0.155 |
| NSAID medication <sup>∘</sup> | 39 (24%) | 434 (26%) | 473 (26%) | 0.062 |
| Tramadol medication | <11 | 132 (8.0%) | -- | 0.129 |
| Neuropathic pain medication <sup>∘</sup> | 15 (9.1%) | 141 (8.5%) | 156 (8.6%) | 0.019 |
| Interventions in the 12 months prior to the procedure |  |  |  |  |
| Oral corticosteroids <sup>∘</sup> | 28 (17%) | 270 (16%) | 298 (16%) | 0.016 |
| Corticosteroid injection <sup>∘</sup> | 65 (39%) | 722 (44%) | 787 (43%) | 0.089 |
| Number of corticosteroid injections |  |  |  | 0.159 |
| Mean (SD) | 0.6 (1.0) | 0.8 (1.2) | 0.8 (1.2) |  |
| Median (Q1, Q3) | 0.0 (0.0, 1.0) | 0.0 (0.0, 1.0) | 0.0 (0.0, 1.0) |  |
| Hyaluronic acid injection | 23 (14%) | 176 (11%) | 199 (11%) | 0.100 |
| Number of hyaluronic acid injections |  |  |  | 0.063 |
| Mean (SD) | 0.3 (0.7) | 0.2 (0.7) | 0.2 (0.7) |  |
| Median (Q1, Q3) | 0.0 (0.0, 0.0) | 0.0 (0.0, 0.0) | 0.0 (0.0, 0.0) |  |
| Physical therapy | 74 (45%) | 636 (39%) | 710 (39%) | 0.128 |
| Number of PT visits |  |  |  | 0.268 |
| Mean (SD) | 5.1 (10.0) | 2.7 (7.0) | 3.0 (7.4) |  |
| Median (Q1, Q3) | 0.0 (0.0, 6.0) | 0.0 (0.0, 2.0) | 0.0 (0.0, 2.0) |  |
| Health seeking behavior in the 12 months prior to the procedure |  |  |  |  |
| Outpatient visit | 165 (100%) | 1,650 (100%) | 1,815 (100%) | 0.000 |
| Number of outpatient visits |  |  |  | 0.134 |
| Mean (SD) | 20.9 (16.6) | 18.8 (13.8) | 19.0 (14.1) |  |
| Median (Q1, Q3) | 16.0 (9.0, 27.0) | 15.0 (10.0, 23.0) | 15.0 (10.0, 24.0) |  |
| ER visit | 21 (13%) | 187 (11%) | 208 (11%) | 0.043 |
| Number of ER visits |  |  |  | 0.041 |
| Mean (SD) | 0.1 (0.4) | 0.2 (0.5) | 0.2 (0.5) |  |
| Median (Q1, Q3) | 0.0 (0.0, 0.0) | 0.0 (0.0, 0.0) | 0.0 (0.0, 0.0) |  |
| Chiropractic visit <sup>∘</sup> | 35 (21%) | 353 (21%) | 388 (21%) | 0.004 |
| Number of chiropractic visits |  |  |  | 0.067 |
| Mean (SD) | 2.2 (5.7) | 1.8 (5.1) | 1.9 (5.1) |  |
| Median (Q1, Q3) | 0.0 (0.0, 0.0) | 0.0 (0.0, 0.0) | 0.0 (0.0, 0.0) |  |
| Knee x-ray | 116 (70%) | 1,538 (93%) | 1,654 (91%) | NA* |
| MRI of lower joint | 98 (59%) | 689 (42%) | 787 (43%) | NA* |
| Knee x-ray or MRI of lower joint | 145 (88%) | 1,570 (95%) | 1,715 (94%) | NA* |
| Other severity proxies in the 12 months prior to the procedure |  |  |  |  |
| Osteoarthritis outpatient visit for the treated knee | 165 (100%) | 1,650 (100%) | 1,815 (100%) | 0.000 |
| Number of osteoarthritis outpatient visits <sup>∘</sup> |  |  |  | 0.089 |
| Mean (SD) | 4.1 (3.4) | 4.3 (2.6) | 4.3 (2.7) |  |
| Median (Q1, Q3) | 3.0 (2.0, 5.0) | 4.0 (3.0, 5.0) | 4.0 (2.0, 5.0) |  |
| Number of OA outpatient visits for the treated knee |  |  |  | 0.146 |
| Mean (SD) | 3.6 (2.7) | 4.0 (2.4) | 4.0 (2.5) |  |
| Median (Q1, Q3) | 3.0 (2.0, 5.0) | 3.0 (2.0, 5.0) | 3.0 (2.0, 5.0) |  |
| Outpatient visit with an 'M' dx code | 165 (100%) | 1,650 (100%) | 1,815 (100%) | 0.000 |
| Number of outpatient visits with an 'M' dx code |  |  |  | 0.154 |
| Mean (SD) | 11.4 (11.6) | 9.8 (9.4) | 10.0 (9.7) |  |
| Median (Q1, Q3) | 8.0 (4.0, 14.0) | 7.0 (4.0, 12.0) | 7.0 (4.0, 12.0) |  |

<sup>∘</sup> denotes a confounder that was included in propensity score matching – other confounders were balanced prior to matching NA\* denotes variables that are bundled with surgery (likely included in diagnostic work up or necessary for meniscectomy reimbursement) or Regenexx (treating both knees is common even when one knee is primary) and therefore not regarded as confounders and SMDs are not reported

**Table 3.** Post-matching demographic and clinical characteristics – PRP versus Meniscectomy.

| Characteristic | PRP<br>N = 167 | Meniscectomy<br>N = 1,670 | Overall<br>N = 1,837 | ASMD |
| --- | --- | --- | --- | --- |
| Continuous commercial health plan enrollment after index procedure |  |  |  |  |
| CE for 12 months | 167 (100%) | 1,670 (100%) | 1,837 (100%) | 0.000 |
| CE for 24 months | 105 (63%) | 1,224 (73%) | 1,329 (72%) | 0.225 |
| CE for 36 months | 63 (38%) | 842 (50%) | 905 (49%) | 0.258 |
| CE for 48 months | 35 (21%) | 550 (33%) | 585 (32%) | 0.272 |
| Location |  |  |  | NA* |
| Right | 60 (36%) | 848 (51%) | 908 (49%) |  |
| Left | 63 (38%) | 822 (49%) | 885 (48%) |  |
| Both | 44 (26%) | -- | 44 (2.4%) |  |
| Sex |  |  |  | 0.118 |
| Female | 80 (48%) | 702 (42%) | 782 (43%) |  |
| Male | 87 (52%) | 968 (58%) | 1,055 (57%) |  |
| Age (years) |  |  |  | 0.078 |
| Mean (SD) | 51.6 (11.0) | 50.7 (11.3) | 50.8 (11.3) |  |
| Median (Q1, Q3) | 54.0 (46.0, 59.0) | 53.0 (46.0, 59.0) | 53.0 (46.0, 59.0) |  |
| Geographic region |  |  |  | 0.093 |
| Northeast | 39 (23%) | 445 (27%) | 484 (26%) |  |
| Midwest | 69 (41%) | 675 (40%) | 744 (41%) |  |
| South | 37 (22%) | 380 (23%) | 417 (23%) |  |
| West | 22 (13%) | 170 (10%) | 192 (11%) |  |
| Injury to the treated knee within 12 months prior to the procedure |  |  |  |  |
| Meniscus tear | 23 (14%) | 1,327 (79%) | 1,350 (73%) | NA* |
| Other knee injuries (cartilage tear, subluxations/<br>dislocations, sprains) | <11 | 108 (6.5%) | -- | 0.236 |
| Orthopedic diagnoses on the treated knee within 12 months prior to the procedure |  |  |  |  |
| Osteoarthritis<sup>∘</sup> | 48 (29%) | 551 (33%) | 599 (33%) | 0.092 |
| Acquired deformity of the knee | <11 | <11 | -- | 0.049 |
| Chondromalacia patellae | 30 (18%) | 293 (18%) | 323 (18%) | 0.011 |
| Other disorders of patella | <11 | <11 | -- | 0.071 |
| Derangement of meniscus | 14 (8.4%) | 415 (25%) | 429 (23%) | NA* |
| Derangement, unspecified | <11 | 144 (8.6%) | -- | NA* |
| Derangement, instability | <11 | 58 (3.5%) | -- | NA* |
| Severe derangement | <11 | 61 (3.7%) | -- | NA* |
| Spontaneous disruption of ligaments | <11 | <11 | -- | 0.000 |
| Musculoskeletal conditions (not exclusive to the treated knee) within 12 months prior to the procedure |  |  |  |  |
| Knee effusion<sup>∘</sup> | 23 (14%) | 249 (15%) | 272 (15%) | 0.032 |
| Number of knee effusions |  |  |  | 0.104 |
| Mean (SD) | 0.2 (0.6) | 0.2 (0.8) | 0.2 (0.7) |  |
| Median (Q1, Q3) | 0.0 (0.0, 0.0) | 0.0 (0.0, 0.0) | 0.0 (0.0, 0.0) |  |
| Number of knee effusions among patients who had at least one |  |  |  | 0.104 |
| Mean (SD) | 1.5 (0.6) | 1.4 (1.5) | 1.4 (1.5) |  |
| Median (Q1, Q3) | 1.0 (1.0, 2.0) | 1.0 (1.0, 1.0) | 1.0 (1.0, 1.0) |  |
| Missing | 144 | 1,421 | 1,565 |  |
| Enthesopathy of the lower limb, excluding foot | 21 (13%) | 115 (6.9%) | 136 (7.4%) | 0.193 |
| Other knee joint disorders<sup>∘</sup> | 103 (62%) | 1,102 (66%) | 1,205 (66%) | 0.090 |
| Disorders of synovium and tendon | <11 | 64 (3.8%) | -- | 0.123 |
| Other soft tissue disorders<sup>∘</sup> | 76 (46%) | 803 (48%) | 879 (48%) | 0.052 |
| Biomechanical lesions | <11 | 27 (1.6%) | -- | 0.092 |
| Dorsopathy<sup>∘</sup> | 76 (46%) | 796 (48%) | 872 (47%) | 0.043 |
| Osteoarthritis elsewhere | 34 (20%) | 265 (16%) | 299 (16%) | 0.117 |
| Injury to other parts of the body | 46 (28%) | 427 (26%) | 473 (26%) | 0.045 |
| Injury to adjacent parts of the body | 17 (10%) | 192 (11%) | 209 (11%) | 0.042 |
| General comorbidities documented in the 12 months prior to the procedure |  |  |  |  |
| Obesity | 22 (13%) | 225 (13%) | 247 (13%) | 0.009 |
| Diabetes | 16 (9.6%) | 162 (9.7%) | 178 (9.7%) | 0.004 |
| Depression/Anxiety/Bipolar | 26 (16%) | 271 (16%) | 297 (16%) | 0.018 |
| Medications within 12 months prior to the procedure |  |  |  |  |
| Opioid medication<sup>∘</sup> | 32 (19%) | 290 (17%) | 322 (18%) | 0.047 |
| NSAID medication <sup>∘</sup> | 31 (19%) | 258 (15%) | 289 (16%) | 0.083 |
| Tramadol medication | <11 | 48 (2.9%) | -- | 0.100 |
| Neuropathic pain medication | 16 (9.6%) | 106 (6.3%) | 122 (6.6%) | 0.120 |
| Interventions in the 12 months prior to the procedure |  |  |  |  |
| Oral corticosteroids <sup>∘</sup> | 26 (16%) | 257 (15%) | 283 (15%) | 0.005 |
| Corticosteroid injection <sup>∘</sup> | 41 (25%) | 373 (22%) | 414 (23%) | 0.052 |
| Number of corticosteroid injections |  |  |  | 0.016 |
| Mean (SD) | 0.4 (0.8) | 0.4 (0.9) | 0.4 (0.9) |  |
| Median (Q1, Q3) | 0.0 (0.0, 0.0) | 0.0 (0.0, 0.0) | 0.0 (0.0, 0.0) |  |
| Hyaluronic acid injection | <11 | <11 | -- | 0.079 |
| Number of hyaluronic acid injections |  |  |  | 0.092 |
| Mean (SD) | 0.0 (0.2) | 0.0 (0.1) | 0.0 (0.1) |  |
| Median (Q1, Q3) | 0.0 (0.0, 0.0) | 0.0 (0.0, 0.0) | 0.0 (0.0, 0.0) |  |
| Physical therapy <sup>∘</sup> | 70 (42%) | 710 (43%) | 780 (42%) | 0.012 |
| Number of PT visits |  |  |  | 0.104 |
| Mean (SD) | 5.6 (14.3) | 4.3 (9.1) | 4.4 (9.6) |  |
| Median (Q1, Q3) | 0.0 (0.0, 6.0) | 0.0 (0.0, 5.0) | 0.0 (0.0, 5.0) |  |
| Health seeking behavior in the 12 months prior to the procedure |  |  |  |  |
| Outpatient visit | 166 (99%) | 1,668 (100%) | 1,834 (100%) | 0.080 |
| Number of outpatient visits |  |  |  | 0.013 |
| Mean (SD) | 18.7 (17.3) | 18.9 (15.8) | 18.9 (16.0) |  |
| Median (Q1, Q3) | 14.0 (8.0, 24.0) | 14.0 (8.0, 25.0) | 14.0 (8.0, 25.0) |  |
| ER visit | 18 (11%) | 206 (12%) | 224 (12%) | 0.049 |
| Number of ER visits |  |  |  | 0.081 |
| Mean (SD) | 0.1 (0.4) | 0.2 (0.6) | 0.2 (0.6) |  |
| Median (Q1, Q3) | 0.0 (0.0, 0.0) | 0.0 (0.0, 0.0) | 0.0 (0.0, 0.0) |  |
| Chiropractic visit <sup>∘</sup> | 37 (22%) | 392 (23%) | 429 (23%) | 0.031 |
| Number of chiropractic visits |  |  |  | 0.046 |
| Mean (SD) | 3.1 (13.2) | 2.6 (7.2) | 2.6 (8.0) |  |
| Median (Q1, Q3) | 0.0 (0.0, 0.0) | 0.0 (0.0, 0.0) | 0.0 (0.0, 0.0) |  |
| Knee x-ray | 59 (35%) | 1,365 (82%) | 1,424 (78%) | NA* |
| MRI of lower joint | 54 (32%) | 1,338 (80%) | 1,392 (76%) | NA* |
| Knee x-ray or MRI of lower joint | 79 (47%) | 1,527 (91%) | 1,606 (87%) | NA* |
| Other severity proxies in the 12 months prior to the procedure |  |  |  |  |
| Osteoarthritis outpatient visit <sup>∘</sup> | 73 (44%) | 786 (47%) | 859 (47%) | 0.067 |
| Number of osteoarthritis outpatient visits |  |  |  | 0.069 |
| Mean (SD) | 0.8 (1.6) | 0.7 (1.7) | 0.7 (1.6) |  |
| Median (Q1, Q3) | 0.0 (0.0, 1.0) | 0.0 (0.0, 1.0) | 0.0 (0.0, 1.0) |  |
| Number of OA outpatient visits for the treated knee | 48 (29%) | 551 (33%) | 599 (33%) | 0.092 |
| Outpatient visit with an 'M' dx code | 160 (96%) | 1,618 (97%) | 1,778 (97%) | 0.057 |
| Number of outpatient visits with an 'M' dx code <sup>∘</sup> |  |  |  | 0.003 |
| Mean (SD) | 9.4 (10.6) | 9.4 (11.5) | 9.4 (11.4) |  |
| Median (Q1, Q3) | 5.0 (2.0, 12.0) | 5.0 (2.0, 12.0) | 5.0 (2.0, 12.0) |  |

<sup>∘</sup> denotes a confounder that was included in propensity score matching – other confounders were balanced prior to matching NA\* denotes variables that are bundled with surgery (likely included in diagnostic work up or necessary for meniscectomy reimbursement) or Regenexx (treating both knees is common even when one knee is primary) and therefore not regarded as confounders and SMDs are not reported

### Health care use

#### BMAC with or without PRP versus TKA/pKA

After BMAC/PRP, the rates of post-procedure TKA/pKA were below the cell suppression limit of <11 events at 12 and 24 months among those continuously enrolled during those follow-up timeframes (Table 4 & 5). Rates of post-procedure BMAC/PRP or PRP re-injection, i.e., booster treatments on the same knee, were 8.5% at 12 months and 14.9% at 24 months. We observed lower rates (± SD) of outpatient visits (4.6 ± 5.0 vs 7.2 ± 7.5, p<0.001), physical therapy visits (10.1 ± 13.9 vs 15.9 ± 14.8, p<0.001), knee x-ray (15% vs 94%, p<0.001), and opioid use (35% vs 44%, p=0.02) among those receiving BMAC/PRP versus TKA/pKA, but a higher rate of MRI of the lower joints (13% vs 4.3%, p<0.001) at 12 months. Rates of outpatient visits (7.6 ± 6.8 vs 10.9 ± 10.6, p<0.001), physical therapy visits (16.6 ± 23.4 vs 17.9 ± 17.9, p=0.003) and knee x-ray (28% vs 96%, p<0.001) were also lower among those receiving BMAC/PRP at 24 months, but use of MRI was lower (23% vs 7.7%, p<0.001) among those undergoing TKA/pKA. Rates of all other types of health care use were not different between groups.

**Table 4.** Healthcare Resource Utilization 12 months post procedure – BMAC with or without PRP versus TKA/pKA.

| Characteristic | BMAC ± PRP<br>N = 165 | TKA/pKA<br>N = 1,650 | Overall<br>N = 1,815 | p-value |
| --- | --- | --- | --- | --- |
| Number of outpatient visits |  |  |  | < 0.001 |
| Mean (SD) | 4.6 (5.0) | 7.2 (7.5) | 7.0 (7.4) |  |
| Median (Q1, Q3) | 3.0 (1.0, 6.0) | 5.0 (3.0, 9.0) | 5.0 (3.0, 9.0) |  |
| Number of physical therapy visits |  |  |  | < 0.001 |
| Mean (SD) | 10.1 (13.9) | 15.9 (14.8) | 15.4 (14.8) |  |
| Median (Q1, Q3) | 5.0 (0.0, 14.0) | 13.0 (4.0, 23.0) | 12.0 (3.0, 23.0) |  |
| Knee x-ray | 24 (14.5%) | 1,559 (94.5%) | 1,583 (87.2%) | < 0.001 |
| MRI of lower joint | 21 (12.7%) | 71 (4.3%) | 92 (5.1%) | < 0.001 |
| CT of lower extremity | <11 | 65 (3.9%) | -- | 0.25 |
| Subsequent same side TKA/pKA | <11 | 187 (11.3%) | -- | 0.007 |
| Subsequent same side Meniscectomy | <11 | <11 | -- | -- |
| Subsequent same side ACL/MCL procedure | <11 | <11 | -- | -- |
| Subsequent orthobiologic procedure - any knee | 22 (13.3%) | -- | -- | -- |
| Subsequent orthobiologic procedure – same knee | 14 (8.5%) | -- | -- | -- |
| Deep vein thrombosis inpatient care | <11 | <11 | -- | -- |
| Hematoma inpatient care | <11 | <11 | -- | -- |
| Infection inpatient care | <11 | 14 (0.8%) | -- | -- |
| Pulmonary embolism inpatient care | <11 | <11 | -- | -- |
| Other post-surgical embolism inpatient care | <11 | <11 | -- | -- |
| DVT, PE, or OE inpatient care | <11 | 12 (0.7%) | -- | -- |
| Opioid medication | 57 (34.5%) | 730 (44.2%) | 787 (43.4%) | 0.02 |

**Table 5.** Healthcare Resource Utilization 24 months post procedure – BMAC with or without PRP versus TKA/pKA.

| Characteristic | BMAC ± PRP<br>N = 114 | TKA/pKA<br>N = 1,111 | Overall<br>N = 1,225 | p-value |
| --- | --- | --- | --- | --- |
| Number of outpatient visits |  |  |  | < 0.001 |
| Mean (SD) | 7.6 (6.8) | 10.9 (10.6) | 10.6 (10.3) |  |
| Median (Q1, Q3) | 5.0 (2.0, 11.0) | 8.0 (4.0, 14.0) | 8.0 (4.0, 14.0) |  |
| Number of physical therapy visits |  |  |  | 0.003 |
| Mean (SD) | 16.6 (23.4) | 17.9 (17.9) | 17.8 (18.5) |  |
| Median (Q1, Q3) | 7.5 (0.0, 25.0) | 14.0 (4.0, 26.0) | 14.0 (4.0, 25.0) |  |
| Knee x-ray | 32 (28.1%) | 1,063 (95.7%) | 1,095 (89.4%) | < 0.001 |
| MRI of lower joint | 26 (22.8%) | 86 (7.7%) | 112 (9.1%) | < 0.001 |
| CT of lower extremity | <11 | 70 (6.3%) | -- | 0.82 |
| Subsequent same side TKA/pKA | <11 | 136 (12.2%) | -- | 0.22 |
| Subsequent same side Meniscectomy | <11 | <11 | -- | -- |
| Subsequent same side ACL/MCL procedure | <11 | <11 | -- | -- |
| Subsequent orthobiologic procedure - any knee | 20 (17.5%) | -- | -- | -- |
| Subsequent orthobiologic procedure – same knee | 17 (14.9%) | -- | -- | -- |
| Deep vein thrombosis inpatient care | <11 | <11 | -- | -- |
| Hematoma inpatient care | <11 | <11 | -- | -- |
| Infection inpatient care | <11 | 18 (1.6%) | -- | -- |
| Pulmonary embolism inpatient care | <11 | <11 | -- | -- |
| Other post-surgical embolism inpatient care | <11 | <11 | -- | -- |
| DVT, PE, or OE inpatient care | <11 | 16 (1.4%) | -- | -- |
| Opioid medication | 48 (42.1%) | 516 (46.4%) | 564 (46.0%) | 0.43 |

**Table 6:** Healthcare Resource Utilization 12 months post procedure – PRP versus Meniscectomy.

| Characteristic | PRP<br>N = 167 | Meniscectomy<br>N = 1,670 | Overall<br>N = 1,837 | p-value |
| --- | --- | --- | --- | --- |
| Number of outpatient visits |  |  |  | 0.15 |
| Mean (SD) | 4.2 (5.5) | 3.8 (5.5) | 3.9 (5.5) |  |
| Median (Q1, Q3) | 2.0 (1.0, 5.0) | 2.0 (0.0, 5.0) | 2.0 (0.0, 5.0) |  |
| Number of physical therapy visits |  |  |  | 0.003 |
| Mean (SD) | 6.5 (14.4) | 7.0 (10.4) | 6.9 (10.8) |  |
| Median (Q1, Q3) | 0.0 (0.0, 9.0) | 2.0 (0.0, 10.0) | 2.0 (0.0, 10.0) |  |
| Knee x-ray | 23 (13.8%) | 298 (17.8%) | 321 (17.5%) | 0.22 |
| MRI of lower joint | 14 (8.4%) | 175 (10.5%) | 189 (10.3%) | 0.47 |
| CT of lower extremity | <11 | 14 (0.8%) | -- | -- |
| Subsequent same side TKA/PKA | <11 | 32 (1.9%) | -- | -- |
| Subsequent same side Meniscectomy | <11 | 85 (5.1%) | -- | 0.09 |
| Subsequent same side ACL/MCL procedure | <11 | <11 | -- | -- |
| Subsequent orthobiologic procedure - any knee | 20 (12.0%) | -- | -- | -- |
| Subsequent orthobiologic procedure – same knee | 17 (10.2%) | -- | -- | -- |
| Deep vein thrombosis inpatient care | <11 | <11 | -- | -- |
| Hematoma inpatient care | <11 | <11 | -- | -- |
| Infection inpatient care | <11 | <11 | -- | -- |
| Pulmonary embolism inpatient care | <11 | <11 | -- | -- |
| Other post-surgical embolism inpatient care | <11 | <11 | -- | -- |
| DVT, PE, or OE inpatient care | <11 | <11 | -- | -- |
| Opioid medication | 45 (26.9%) | 218 (13.1%) | 263 (14.3%) | < 0.001 |

**Table 7.**
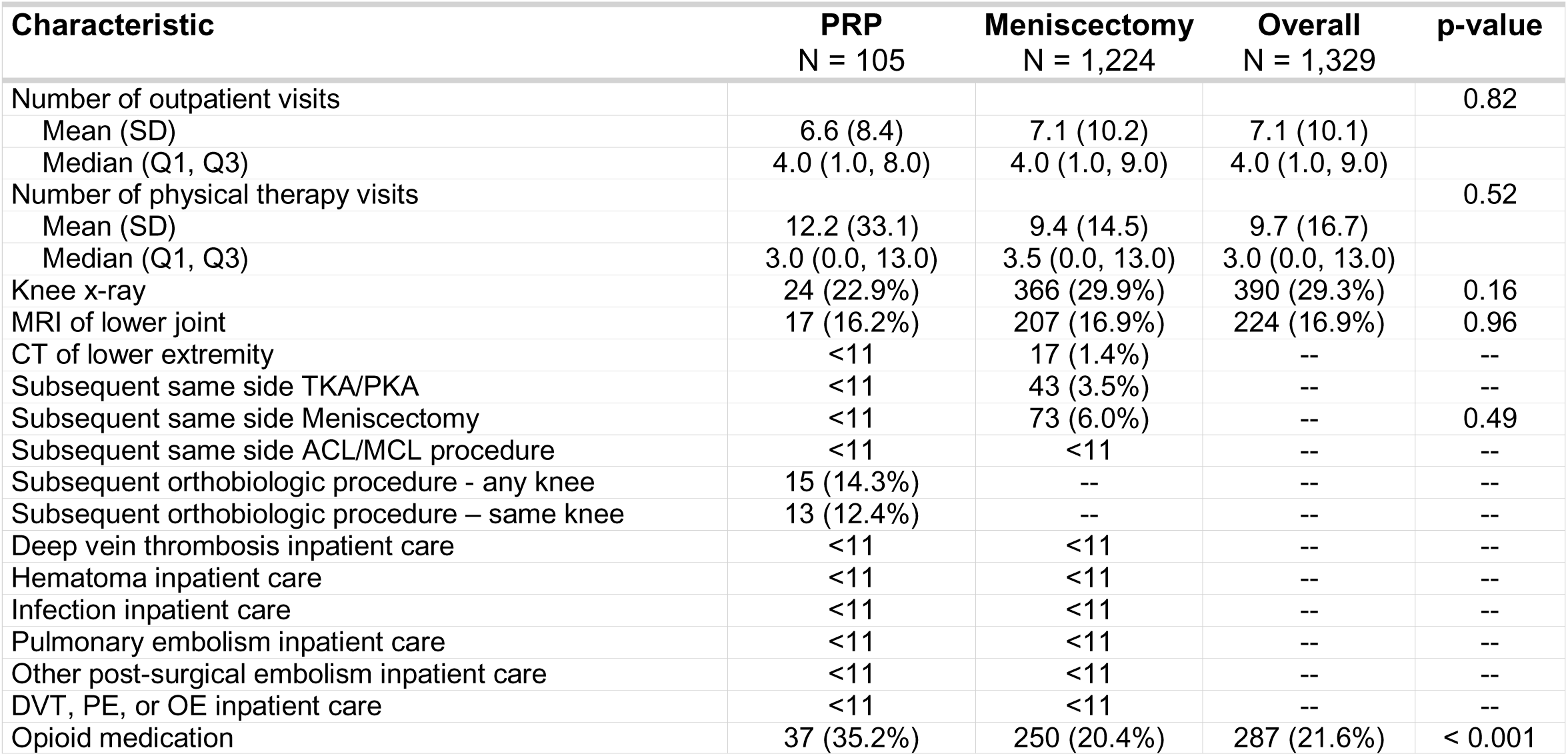
Healthcare Resource Utilization 24 months post procedure – PRP versus Meniscectomy.

| Characteristic | PRP<br>N = 105 | Meniscectomy<br>N = 1,224 | Overall<br>N = 1,329 | p-value |
| --- | --- | --- | --- | --- |
| Number of outpatient visits |  |  |  | 0.82 |
| Mean (SD) | 6.6 (8.4) | 7.1 (10.2) | 7.1 (10.1) |  |
| Median (Q1, Q3) | 4.0 (1.0, 8.0) | 4.0 (1.0, 9.0) | 4.0 (1.0, 9.0) |  |
| Number of physical therapy visits |  |  |  | 0.52 |
| Mean (SD) | 12.2 (33.1) | 9.4 (14.5) | 9.7 (16.7) |  |
| Median (Q1, Q3) | 3.0 (0.0, 13.0) | 3.5 (0.0, 13.0) | 3.0 (0.0, 13.0) |  |
| Knee x-ray | 24 (22.9%) | 366 (29.9%) | 390 (29.3%) | 0.16 |
| MRI of lower joint | 17 (16.2%) | 207 (16.9%) | 224 (16.9%) | 0.96 |
| CT of lower extremity | <11 | 17 (1.4%) | -- | -- |
| Subsequent same side TKA/PKA | <11 | 43 (3.5%) | -- | -- |
| Subsequent same side Meniscectomy | <11 | 73 (6.0%) | -- | 0.49 |
| Subsequent same side ACL/MCL procedure | <11 | <11 | -- | -- |
| Subsequent orthobiologic procedure - any knee | 15 (14.3%) | -- | -- | -- |
| Subsequent orthobiologic procedure – same knee | 13 (12.4%) | -- | -- | -- |
| Deep vein thrombosis inpatient care | <11 | <11 | -- | -- |
| Hematoma inpatient care | <11 | <11 | -- | -- |
| Infection inpatient care | <11 | <11 | -- | -- |
| Pulmonary embolism inpatient care | <11 | <11 | -- | -- |
| Other post-surgical embolism inpatient care | <11 | <11 | -- | -- |
| DVT, PE, or OE inpatient care | <11 | <11 | -- | -- |
| Opioid medication | 37 (35.2%) | 250 (20.4%) | 287 (21.6%) | < 0.001 |

#### PRP versus Meniscectomy

Rates of meniscectomy following PRP were below the cell suppression limit of <11 events at 12 and 24 months (Tables 6 & 7). Rates of post-procedure BMAC/PRP or PRP re-injection on the same knee, i.e., booster treatment, were 10% at 12 months and 12% at 24 months. Number of physical therapy visits was lower following PRP compared to meniscectomy (6.5 ± 14.4 vs 7.0 ± 10.4, p=0.003) at 12 months but not at 24 months. Opioid medication use was higher in those who received PRP compared to meniscectomy at 12 months (27% vs 13%) and 24 months (35% vs 20%). We observed no other significant differences in HCRU between the two treatments at 12 months and 24 months.

### Costs

#### BMAC with or without PRP versus TKA/pKA

At 12 months, estimated mean difference (95% CI) between BMAC/PRP versus TKA/pKA ranged from $13,556 (12,394, 14,718; p-value<0.001) using commercial payer costs, to $39,913 (37,900, 41,926; p-value<0.001) using external Medicare costs × 2 (Table 8). Surgery was associated with higher costs using all costing estimates.

**Table 8.** Costs – BMAC ± PRP versus TKA/PKA at 12- and 24-months post procedure.

| Characteristic | BMAC ± PRP<br>N = 165 | TKA/PKA<br>N = 1,650 | Estimated mean<br>difference (95% CI) | p-value |
| --- | --- | --- | --- | --- |
| **12 months post-procedure** |  |  |  |  |
| External Medicare costs (\$) | | | | |
| Mean (SD) | 15,195 (6,138) | 28,948 (8,825) | 13,753 (12,720, 14,785) | <0.001 |
| Median (Q1, Q3) | 13,135 (12,285, 14,785) | 25,550 (25,150, 26,750) |  |  |
| External Medicare cost x2 (\$) | | | | |
| Mean (SD) | 17,982 (11,907) | 57,895 (17,650) | 39,913 (37,900, 41,926) | <0.001 |
| Median (Q1, Q3) | 14,285 (12,585, 16,985) | 51,100 (50,300, 53,500) |  |  |
| Commercial payer average costs (\$) | | | | |
| Mean (SD) | 16,699 (6,889) | 30,255 (10,069) | 13,556 (12,394, 14,718) | <0.001 |
| Median (Q1, Q3) | 14,433 (13,013, 16,668) | 26,694 (24,956, 29,961) |  |  |
| Aggregate costing method (\$) | | | | |
| Mean (SD) | 18,759 (11,329) | 37,497 (23,591) | 18,738 (16,662, 20,814) | <0.001 |
| Median (Q1, Q3) | 13,980 (12,857, 18,086) | 32,703 (24,729, 44,701) |  |  |
| Aggregate costing method – truncated (\$) | | | | |
| Mean (SD) | 18,692 (10,758) | 36,094 (16,200) | 17,402 (15,579, 19,226) | <0.001 |
| Median (Q1, Q3) | 13,980 (12,857, 18,086) | 32,703 (24,729, 44,701) |  |  |
| **24 months post-procedure** |  |  |  |  |
|  | **BMAC ± PRP**<br>**N = 114** | **TKA/PKA**<br>**N = 1,111** | **Estimated mean**<br>**difference (95% CI)** | **p-**<br>**value** |
| External (Medicare) costs (\$) | | | | |
| Mean (SD) | 18,286 (9,924) | 30,178 (10,348) | 11,892 (9,961, 13,822) | <0.001 |
| Median (Q1, Q3) | 14,385 (12,635, 17,920) | 26,200 (25,350, 27,950) |  |  |
| External Medicare cost x2 (\$) | | | | |
| Mean (SD) | 23,629 (19,287) | 60,356 (20,695) | 36,727 (32,964, 40,489) | <0.001 |
| Median (Q1, Q3) | 16,477 (13,285, 22,385) | 52,400 (50,700, 55,900) |  |  |
| Commercial payer average costs (\$) | | | | |
| Mean (SD) | 20,968 (11,706) | 32,764 (12,701) | 11,796 (9,510, 14,082) | <0.001 |
| Median (Q1, Q3) | 16,653 (13,553, 23,143) | 28,176 (25,855, 33,178) |  |  |
| Commercial payer actual costs (\$) | | | | |
| Mean (SD) | 27,415 (42,437) | 41,295 (26,696) | 13,881 (5,892, 21,869) | <0.001 |
| Median (Q1, Q3) | 16,170 (13,086, 28,974) | 35,256 (26,378, 48,838) |  |  |
| Commercial payer actual costs – truncated (\$) | | | | |
| Mean (SD) | 23,609 (16,486) | 39,683 (18,658) | 16,074 (12,839, 19,309) | <0.001 |
| Median (Q1, Q3) | 16,170 (13,086, 28,974) | 35,256 (26,378, 48,838) |  |  |

At 24 months, estimated mean difference for BMAC/PRP versus TKA/pKA range from $11,796 (9,510, 14,082; p-value<0.001) using commercial payer costs to $36,727 (32,964, 40,489; p-value<0.001) using external Medicare costs × 2 (Table 8), with surgery associated with higher costs using all costing estimates.

#### PRP versus Meniscectomy

At 12 months, estimated mean difference for PRP versus meniscectomy using Medicare cost estimates was $4 (-672, 681; p=0.991). Meniscectomy was associated with higher costs at 12 months using all other costing estimates (Table 9).

**Table 9.** Costs – PRP versus meniscectomy at 12- and 24-months post procedure.

| Characteristic | PRP<br>N = 167 | Meniscectomy<br>N = 1,670 | Estimated mean<br>difference (95% CI) | p-value |
| --- | --- | --- | --- | --- |
| **12 months post-procedure** |  |  |  |  |
| External (Medicare) costs (\$) | | | | |
| Mean (SD) | 5,256 (4,195) | 5,260 (4,652) | 4 (-672, 681) | 0.991 |
| Median (Q1, Q3) | 3,785 (3,285, 5,585) | 4,023 (3,523, 5,223) |  |  |
| External Medicare cost x2 (\$) | | | | |
| Mean (SD) | 6,629 (6,584) | 10,520 (9,305) | 3,892 (2,794, 4,989) | <0.001 |
| Median (Q1, Q3) | 4,485 (3,485, 6,985) | 8,046 (7,046, 10,446) |  |  |
| Commercial payer average costs (\$) | | | | |
| Mean (SD) | 6,701 (5,559) | 9,279 (5,725) | 2,578 (1,688, 3,468) | <0.001 |
| Median (Q1, Q3) | 4,749 (3,477, 7,092) | 7,523 (6,379, 9,527) |  |  |
| Commercial payer actual costs (\$) | | | | |
| Mean (SD) | 7,580 (8,486) | 9,511 (12,929) | 1,931 (498, 3,364) | 0.008 |
| Median (Q1, Q3) | 4,423 (3,295, 7,020) | 6,565 (4,504, 10,229) |  |  |
| Commercial payer actual costs – truncated (\$) | | | | |
| Mean (SD) | 6,892 (5,865) | 8,428 (5,632) | 1,536 (603, 2,469) | 0.001 |
| Median (Q1, Q3) | 4,423 (3,295, 7,020) | 6,565 (4,504, 10,229) |  |  |
| **24 months post-procedure** |  |  |  |  |
|  | **PRP**<br>**N = 105** | **Meniscectomy**<br>**N = 1,224** | **Estimated mean**<br>**difference (95% CI)** | **p-value** |
| External (Medicare) costs (\$) | | | | |
| Mean (SD) | 5,990 (4,598) | 6,473 (6,900) | 484 (-482, 1,449) | 0.326 |
| Median (Q1, Q3) | 4,485 (3,385, 6,621) | 4,523 (3,723, 6,223) |  |  |
| External Medicare cost x2 (\$) | | | | |
| Mean (SD) | 8,055 (7,946) | 12,946 (13,800) | 4891 (3,178, 6,605) | <0.001 |
| Median (Q1, Q3) | 5,885 (3,685, 9,685) | 9,046 (7,446, 12,446) |  |  |
| Commercial payer average costs (\$) | | | | |
| Mean (SD) | 8,177 (6,457) | 11,534 (8,709) | 3,357 (2,022, 4,691) | <0.001 |
| Median (Q1, Q3) | 5,818 (3,869, 9,632) | 8,739 (6,951, 12,045) |  |  |
| Commercial payer actual costs (\$) | | | | |
| Mean (SD) | 9,414 (9,484) | 12,013 (13,855) | 2,599 (616, 4,582) | 0.01 |
| Median (Q1, Q3) | 6,171 (3,659, 9,407) | 7,581 (4,967, 12,450) |  |  |
| Commercial payer actual costs – truncated (\$) | | | | |
| Mean (SD) | 9,161 (8,565) | 10,882 (8,821) | 1,721 (1, 3,442) | 0.05 |
| Median (Q1, Q3) | 6,171 (3,659, 9,407) | 7,581 (4,967, 12,450) |  |  |

At 24 months, estimated mean difference for PRP versus meniscectomy using Medicare cost estimates was $484 (-482, 1449; p=0.326). Meniscectomy was associated with higher costs at 24 months using all other costing estimates (Table 9).

### Exploratory analyses at 36 and 48 months

Rates of post-procedure TKA/pKA after BMAC/PRP were below the cell suppression limit of <11 events at 36 and 48 months among those continuously enrolled during those follow-up timeframes. Rates of knee x-ray continued to be higher among those undergoing TKA/pKA, while rates of MRI were higher in those receiving BMAC with or without PRP (Appendix J & K). At 36 months, rates of outpatient visits and physical therapy visits were higher in those undergoing meniscectomy, but these rates were not different between treatments at 48 months. At 36 and 48 months, costs were higher among those that underwent TKA/pKA compared to BMAC with or without PRP using all cost estimates (Appendix L).

Rates of post-procedure meniscectomy in the PRP sample were below the cell suppression limit of <11 events at 36 and 48 months among those continuously enrolled during those follow-up timeframes. Rates of all HCRU were similar across treatment groups at 36 and 48 months except for opioid use, which was higher in those receiving PRP at 36 months (Appendix M & N). At 36 months, costs were higher in those receiving meniscectomy using all costing strategies except external (Medicare) costs, which showed no differences between treatments (Appendix O). At 48 months, those who received meniscectomy had higher costs using all costing estimates except the aggregate and truncated aggregate methods, which showed no difference between groups.

### Expanded Analysis

The analysis with reduced exclusion criteria to overcome cell suppression included n=763 undergoing orthobiologic treatment (PRP or BMAC), n=130,466 undergoing TKA/pKA, and n=241,731 undergoing meniscectomy (Appendix P & Q). Among those who received orthobiologic treatment, rates of subsequent meniscectomy were 2.1% at 24 months and below the cell suppression limit of 11 at 12, 36 and 48 months (Appendix R through U). Among those who received orthobiologic treatment, rates of subsequent TKA/pKA were 1.8%, 4.5%, 6.7%, and 8.5% at 12, 24, 36 and 48 months.

## DISCUSSION

This study addresses a clinically relevant question amid increasing interest in effective, less costly interventions for high-impact musculoskeletal conditions. Our findings suggest that BMAC and PRP treatments may represent a less expensive alternative to surgical options for certain patients with osteoarthritis and meniscal tears. Using multiple costing approaches designed to enhance generalizability and test the sensitivity of our findings, BMAC and PRP procedures and associated downstream HCRU were consistently less expensive or comparable to TKA/pKA and meniscectomy. These results challenge assumptions that surgery is inevitable following orthobiologic treatment, as progression to surgery in this cohort was rare across our co-primary endpoints at 12 and 24 months and remained low among those followed for 48 months.

We cannot directly measure cost-effectiveness due to the lack of quality measures. However, low downstream health care use in the BMAC and PRP cohorts likely reflect sustained symptom improvement, an assumption supported by prior clinical trials showing meaningful improvements in pain and function for similar populations.^10^ Although we lacked patient-reported outcomes in this analysis, the relatively low rates of follow-up care in both groups may indicate satisfactory outcomes, consistent with evidence that patients often do not seek additional care when treatment is effective.^24,28^

Cost differences were most pronounced when comparing BMAC with or without PRP to arthroplasty, while PRP and meniscectomy showed smaller differences, largely attributable to the lower cost of meniscectomy relative to TKA/pKA. Indirect costs such as missed work and travel expenses were not captured but cost differences would likely favor BMAC and PRP treatments given the longer recovery associated with surgery, especially TKA/pKA. Adverse events were difficult to quantify due to data suppression for low cell counts, but available data suggest events were low or absent in both groups.

Generally, more invasive procedures are associated with higher risk of adverse events, which should also be considered when evaluating the risks and benefits of surgery versus orthobiologic use.

Of the limited prior studies that compare clinical outcomes among patients undergoing orthobiologic versus surgical treatment, very few compare downstream costs of care. This makes our study both unique and challenging to directly compare with other literature. A prior simulation study showed PRP alone was not cost-effective compared to TKA, but assumed all patients progressed to arthroplasty within one year.^33^ That assumption is not supported by our data or other longitudinal studies.

Pabinger et al.^31^ assessed four-year outcomes in a small cohort (n=37) with moderate to severe knee osteoarthritis undergoing BMAC treatment and found sustained improvements in function over 4 years with no participants progressing to arthroplasty during follow-up. Our study helps address a critical gap in the literature by demonstrating that orthobiologic treatments can result in similar or lower downstream costs compared to common surgical alternatives for certain patients with osteoarthritis or meniscal tears.

Several factors should be considered when interpreting the generalizability of our findings. The propensity-matched cohorts were well-balanced across demographic and health-related variables, yet they represent a specific subset of patients, namely those eligible for surgery who elected orthobiologic treatment. Selection bias is possible, as individuals preferring BMAC or PRP treatment over surgery may differ in expectations or other unmeasured characteristics that might lead to better outcomes with orthobiologic treatment. Importantly, we should also note the mean age of patients with meniscal tears (approximately 51 years) and osteoarthritis (approximately 56 years). These sample characteristics suggest our findings apply primarily to management of degenerative meniscal tears rather than injury-related cases of meniscal tear that more commonly occur in younger adults.

Similarly, these findings are most relevant to the management of osteoarthritis in middle-aged adults, a group in which earlier stages of the disease are more common. Although we used proxy measures like intensity of health care use to approximate osteoarthritis severity, direct measures like Kellgren-Lawrence grade were unavailable, limiting our ability to assess applicability of these findings across the full spectrum of disease severity. Our matching process likely produced cohorts that were similar in severity, but we cannot report the exact distribution within each group. In summary, our analytic sample likely reflected populations for which the benefits of surgery are less certain, and this should be considered when determining how to apply these results to clinical practice.

A few other limitations are worth noting. Like all studies using claims data, we had to make assumptions about how documentation used for payment reflects clinical decision-making. Our analysis was further constrained because BMAC and PRP treatments were billed directly to Regenexx rather than through traditional commercial payer claims. Direct billing data included less detailed diagnostic information for each procedure. For this reason, complete diagnostic data associated with each BMAC and PRP treatment was not available. However, Regenexx employs a comprehensive utilization review process to ensure patients meet diagnostic criteria for orthopedic surgery, in this case TKA/pKA or meniscectomy. This provides reasonable assurance that comparison cohorts had similar diagnoses warranting treatment. A final limitation is the small sample available for comparisons beyond 24 months due to changes in health insurance coverage over time. Although trends in costs and health care use appeared consistent through 48 months, these findings should be interpreted cautiously given the limited long-term data.

## Conclusion

Our results indicate that BMAC/PRP and PRP are associated with lower commercial payer costs compared to TKA/pKA or meniscectomy, respectively, for some individuals whose age or health status makes the benefit of surgery uncertain. In this sample, patients who received BMAC/PRP and PRP rarely required subsequent surgery in the 1 to 2 years following orthobiologic treatment, and this rate remained low in our expanded analysis. These insights should inform shared decision-making and payer strategies, especially given the substantial economic burden of musculoskeletal conditions. Further research that incorporates clinical outcomes and indirect costs is needed, but our results underscore the importance of offering orthobiologic treatment as an option in patient-centered care.

## Data Availability

Data produced for the present study are not publicly available

## Funding

This study was sponsored by Regenexx. The sponsor had no role in the conduct of the analysis or the interpretation of the data, manuscript preparation, or the decision to submit the manuscript for publication. Duke University investigators had full access to all study data and retained final responsibility for the content of the manuscript and the decision to submit for publication. We wish to acknowledge support from the Biostatistics, Epidemiology and Research Design (BERD) Methods Core funded through Grant Award Number UL1TR002553 from the National Center for Advancing Translational Sciences (NCATS), a component of the National Institutes of Health (NIH). The content is solely the responsibility of the authors and does not necessarily represent the official views of the NIH.

## Notes

### Competing Interest Statement

Dr. Centeno is employed by Regenexx. This study was sponsored by Regenexx. The sponsor had no role conduct of the analysis or interpretation of the data, manuscript preparation, or the decision to submit the manuscript for publication. Duke University investigators had full access to all study data and retained final responsibility for the content of the manuscript and the decision to submit for publication.

### Funding Statement

This study was sponsored by Regenexx. The sponsor had no role conduct of the analysis or interpretation of the data, manuscript preparation, or the decision to submit the manuscript for publication. Duke University investigators had full access to all study data and retained final responsibility for the content of the manuscript and the decision to submit for publication. We wish to acknowledge support from the Biostatistics, Epidemiology and Research Design (BERD) Methods Core funded through Grant Award Number UL1TR002553 from the National Center for Advancing Translational Sciences (NCATS), a component of the National Institutes of Health (NIH). The content is solely the responsibility of the authors and does not necessarily represent the official views of the NIH.

### Author Declarations

The Duke University IRB gave ethical approval for this work (Pro00116431).

### Summary of Updates

Updated methodological description of confounders and cost outcomes; Appendix F

