## Appendices for "Comparative Analysis of Health Care Use and Costs for Orthobiologic versus Surgical Treatments in Economically High-Impact Knee Conditions"

**Appendix A. Defining eligibility criterion for elective procedures preceded by a relevant clinical diagnosis**

A procedure was defined as elective and preceded by a relevant clinical diagnosis based on the following:

- Patient had at least one relevant diagnosis in an outpatient setting within 12 months preceding the Procedure. Relevant diagnoses are every S and M code listed in the confounders section (Appendix D)
- If care was provided in the emergency department within 3 months prior to intervention, they must also have one relevant diagnosis in an outpatient setting occurring after the first service date of that ER visit and preceding the Procedure. If a patient had more than one emergency department visit in the prior 3 months, the visit closest to the procedure date must satisfy this requirement.
- If the procedure (surgery) occurs in the hospital (inpatient setting) it occurs on the first service date of the hospitalization.

### Appendix B. Procedure and diagnostic codes used for analysis

**Table 1B.** CPT codes for identifying surgical knee procedures

| | CPT Code |
|---|---|
| Total knee arthroplasty | 27447 |
| Total knee arthroplasty revision | 27486, 27487 |
| Partial knee arthroplasty | 27446 |
| Arthroscopic partial meniscectomy or repair | 29880-29883 |

**Table 2B.** ICD-10 codes for knee injury

| Diagnosis | Side | ICD-10 diagnosis codes |
|---|---|---|
| Tear of meniscus | Right | S83.200, S83.203, S83.206, S83.211, S83.221, S83.231, S83.241, S83.251, S83.261, S83.271, S83.281 |
| | Left | S83.201, S83.204, S83.207, S83.212, S83.222, S83.232, S83.242, S83.252, S83.262, S83.272, S83.282 |
| | Unspecified | S83.202, S83.205, S83.209, S83.219, S83.229, S83.239, S83.249, S83.259, S83.269, S83.279, S83.289 |
| (MCL tear) Sprain of collateral ligament of knee | Right | S83.401, S83.411, S83.421 |
| | Left | S83.402, S83.412, S83.422 |
| | Unspecified | S83.409, S83.419, S83.429 |
| (ACL tear) Sprain cruciate ligament of knee | Right | S83.501, S83.511, S83.521 |
| | Left | S83.502, S83.512, S83.522 |
| | Unspecified | S83.509, S83.519, S83.529 |
| Tear of articular cartilage of knee | Right | S83.31 |
| | Left | S83.32 |
| | Unspecified | S83.30 |
| Subluxation or dislocation of patella | Right | S83.001, S83.004, S83.011, S83.014, S83.091, S83.094 |
| | Left | S83.002, S83.005, S83.012, S83.015, S83.092, S83.095 |
| | Unspecified | S83.003, S83.006, S83.013, S83.016, S83.093, S83.096 |
| Subluxation or dislocation of knee | Right | S83.101, S83.104, S83.111, S83.114, S83.121, S83.124, S83.131, S83.134, S83.141, S83.144, S83.191, S83.194 |
| | Left | S83.102, S83.105, S83.112, S83.115, S83.122, S83.125, S83.132, S83.135, S83.142, S83.145, S83.192, S83.195 |
| | Unspecified | S83.103, S83.106, S83.113, S83.116, S83.123, S83.126, S83.133, S83.136, S83.143, S83.146, S83.193, S83.196 |
| Sprain of the superior tibiofibular joint and ligament | Right | S83.61 |
| | Left | S83.62 |
| | Unspecified | S83.60 |
| Other sprain of knee | Right | S83.8X1, S83.91 |
| | Left | S83.8X2, S83.92 |

| | | |
|---|---|---|
| | Unspecified | S83.8X9, S83.90 |

**Table 3B.** ICD-10 codes for orthopedic diagnoses of the knee

| Diagnosis | Side | ICD-10 diagnosis codes |
|---|---|---|
| Osteoarthritis | Right | M17.11, M17.31 |
| | Left | M17.12, M17.32 |
| | Bilateral | M17.0, M17.2, M17.4 |
| | Unspecified | M17.10, M17.30, M17.5, M17.9 |
| Acquired deformities of the knee | Right | M21.061, M21.161, M21.261 |
| | Left | M21.062, M21.162, M21.262 |
| | Unspecified | M21.069, M21.169, M21.269 |
| Chondromalacia patellae + Patellofemoral disorders + M94.26 (juvenile code used wrong) | Right | M22.41, M22.2X1, M94.261 |
| | Left | M22.42, M22.2X2, M94.262 |
| | Unspecified | M22.40, M22.2X9, M94.269 |
| Other disorders of patella | Right | M22.8X1, M22.01, M22.3X1 |
| | Left | M22.8X2, M22.02, M22.3X2 |
| | Unspecified | M22.8X9, M22.00, M22.3X9 |
| Derangement of meniscus | Right | M23.300, M23.303, M23.306, M23.311, M23.321, M23.331, M23.341, M23.351, M23.361, M23.200, M23.203, M23.206, M23.211, M23.221, M23.231, M23.241, M23.251, M23.261 |
| | Left | M23.301, M23.304, M23.307, M23.312, M23.322, M23.332, M23.342, M23.352, M23.362, M23.201, M23.204, M23.207, M23.212, M23.222, M23.232, M23.242, M23.252, M23.262 |
| | Unspecified | M23.302, M23.305, M23.309, M23.319, M23.329, M23.339, M23.349, M23.359, M23.369, M23.202, M23.205, M23.209, M23.219, M23.229, M23.239, M23.249, M23.259, M23.269 |
| Derangement – instability | Right | M23.8X1, M23.51, M23.601, M23.611, M23.621, M23.631, M23.641, M23.671 |
| | Left | M23.8X2, M23.52, M23.602, M23.612, M23.622, M23.632, M23.642, M23.672 |
| | Unspecified | M23.8X9, M23.50, M23.609, M23.619, M23.629, M23.639, M23.649, M23.679 |
| More severe internal derangements of knee | Right | M23.41, M23.000, M23.003, M23.006, M23.011, M23.021, M23.031, M23.041, M23.051, M23.061 |
| | Left | M23.42, M23.001, M23.004, M23.007, M23.012, M23.022, M23.032, M23.042, M23.052, M23.062 |
| | Unspecified | M23.40, M23.002, M23.005, M23.009, M23.019, M23.029, M23.039, M23.049, M23.059, M23.069 |
| Derangement – unspecified | Right | M23.91 |
| | Left | M23.92 |
| | Unspecified | M23.90 |

**Appendix C. Ineligible diagnoses**

- Inflammatory syndrome diagnosis in the last 12 months (M04 - M14). Examples:
    - Autoinflammatory syndromes (M04)
    - Rheumatoid arthritis (M06)
    - Unspecified arthropathy (M12)

- Ineligible diagnoses within the 12 months prior to the procedure
    - Connective tissue disorders
        - Systemic connective tissue disorders (M30-M36)
        - Other disorders of the musculoskeletal system and connective tissue (M95)
    - Joint disorders
        - Hemarthrosis (M25.06)
        - Fistula (M25.16)
        - Flail joint (M25.26)
    - Musculoskeletal injuries
        - Open wound of knee and lower leg (S81)
        - Injury of nerves at lower leg level (S84)
        - Injury of blood vessels at lower leg level (S85)
        - Crushing injury of lower leg (S87)
        - Traumatic amputation of lower leg (S88)
    - Infectious arthropathies (M00-M02)
    - Cancer: malignant neoplasms of bone and articular cartilage (C40, C41)

### Appendix D. Definitions and identification of confounders used in analyses

- Demographics
  - Geographic region
  - Age
  - Sex
- Injury to the treated knee within 12 months prior to the procedure
  - Tear of meniscus (S83.2)
    - Note that codes for unspecified knee will be assumed to apply to the treated knee.
  - (MCL tear) Sprain of collateral ligament of knee (S83.4)
  - (ACL tear) Sprain cruciate ligament of knee (S83.5)
  - Other knee injury
    - Tear of articular cartilage of knee (S83.3)
    - Subluxation or dislocation of patella (S83.0)
    - Subluxation or dislocation of knee (S83.1)
    - Sprain of the superior tibiofibular joint and ligament (S83.6)
    - Other sprain of knee (S83.8, S83.9)
- Orthopedic diagnoses on the treated knee within 12 months prior to the procedure
  - Osteoarthritis of the treated knee (M17 codes)
    - Bilateral codes count as a diagnosis on the treated knee, regardless of which knee was treated
  - Acquired deformities of the limb (M21)
  - Disorders of the patella (M22). The following were included separately:
    - Chondromalacia patellae (M22.4) + Patellofemoral disorders (M22.2) + M94.26
    - Other disorders of the patella combined (M22.8, M22.0, M22.3)
  - Derangement (M23 category). The following to be included separately:
    - Meniscus:
      - Derangement of meniscus (M23.3)
      - Derangement of meniscus due to old injury (M23.2)
    - Instability
      - Other internal derangements (M23.8)
      - Chronic instability (M23.5)
      - Spontaneous disruption of ligaments (M23.6)
    - More severe internal derangements of knee including
      - Loose body in the knee (M23.4)
      - Cystic meniscus (M23.0)
    - Unspecified internal derangement (M23.9)
- Musculoskeletal conditions (not exclusive to the treated knee) within 12 months prior to procedure
  - Knee joint disorders (M25 category). The following were included separately:
    - Effusion (M25.46)
    - Other knee joint disorders combined:
      - Pain in knee
        - M25.56
        - Other knee bursopathies
          - M71.06, M71.16, M71.2, M71.46, M71.56, M71.86
      - Stiffness of knee (M25.66)
      - Other instability (M25.36)
      - Osteophyte (M25.76)

- Enthesopathies, lower limb, excluding foot (M76)
- Disorders of synovium and tendon to be combined (N=60). Only including disorders of the knee or lower leg
  - Synovitis and tenosynovitis
    - Lower leg: M65.06, M65.26, M65.86, M65.96
    - Knee: M65.16
  - Spontaneous rupture of synovium and tendon
    - Lower leg: M66.26, M66.36, M66.86
  - Other disorders of synovium and tendon
    - Lower leg: M67.26, M67.96
    - Knee: M67.36, M67.46, M67.5, M67.86
- Other soft tissue disorders generating discomfort
  - Soft tissue disorders related to use, overuse and pressure (M70)
  - Fibroblastic disorders (M72)
  - Shoulder lesions (M75)
  - Other enthesopathies (M77)
  - Other and unspecified soft tissue disorders, not elsewhere classified (M79)
- Biomechanical lesions; segmental and somatic dysfunction of lower extremity (M99.06)
- Dorsopathy. The following to be included as a single group.
  - Deforming dorsopathy (M40-M43)
  - Spondylapathy (M45-M49)
  - Other dorsopathy (M50-M54)
- Osteoarthritis elsewhere. The following were included as a single group.
  - Polyosteoarthritis (M15)
  - Osteoarthritis of the hip (M16)
  - Other osteoarthritis (M18 and M19)
- Injury to other parts of the body (All "S" codes except S80-S89)
- Injury to adjacent parts of the body (S70-S79, S86, S90-S99)

- General comorbidity diagnosis in the 12 months prior to procedure
  - Obesity (E66.0, E66.1, E66.2, E66.8, E66.9, Z68.3, Z68.4, Z68.54, Z68.55, Z68.56)
  - Diabetes
    - Codes: E08 – E11, E13
  - Anxiety/Depression/Bipolar
    - Depression codes: F32, F33
    - Anxiety codes: F41
    - Bipolar codes: F31
- Medications within 12 months prior to the procedure
  - Prescription pain relievers included separately
    - Opioids
    - Prescription NSAIDs
    - Tramadol
    - Neuropathic pain meds (amitriptyline, duloxetine, pregabalin, gabapentin, venlafaxine)
- Intervention history in the 12 months prior to procedure (enumerated)
  - Use of oral corticosteroids
  - Corticosteroid injections (any location)
  - Hyaluronic acid injections (any location)
  - Physical therapy encounters

- Health Seeking behavior / severity in the 12 months prior to procedure
  - # Outpatient medical visits in the past year
  - # ER visits in the prior year
  - # Chiropractic visits in the prior year
  - Knee radiology (Xray)
  - MRI of lower joint
  - Knee radiology (Xray) and/or MRI of lower joint
- Other Severity proxies
  - Severity of osteoarthritis
    - # outpatient encounters with OA diagnosis code of any joint in the 12 months prior to procedure (M15, M16, M17, M18, M19)
  - Severity of osteoarthritis of the treated knee
    - # outpatient visits with a knee OA diagnosis code for the treated knee
  - Severity of orthopedic problems
    - # of outpatient encounters with any M diagnosis code in the in the 12 months prior to procedure

Descriptive data and absolute standardized mean differences (ASMDs) were reported on all the preceding variables. Given the limited sample size it was not possible to put all variables in the propensity score model. The following subset was used in propensity score matching because they were initially imbalanced (not balanced naturally) and not subject to reverse causation (not likely caused by preparation for surgery).

For the TKA/pKA comparison:

- Age
- Meniscus tear on the treated knee (Y/N)
- Derangement of the meniscus on the treated knee (Y/N)
- Effusion, not exclusive to treated knee (Y/N)
- Other knee joint disorder, not exclusive to treated knee (Y/N)
- Obesity (Y/N)
- Diabetes (Y/N)
- NSAID med (Y/N)
- Neuropathic pain med (Y/N)
- Oral corticosteroid (Y/N)
- Corticosteroid injection (Y/N)
- Chiropractic visit (Y/N)
- Number of outpatient encounters with OA diagnosis code of any joint

For the meniscectomy comparison:

- OA on the treated knee (Y/N)
- Effusion, not exclusive to treated knee (Y/N)
- Other knee joint disorder, not exclusive to treated knee (Y/N)
- Other soft tissue disorder, not exclusive to treated knee (Y/N)
- Dorsopathy, not exclusive to treated knee (Y/N)
- Obesity (Y/N)
- Opioid medication (Y/N)
- NSAID med (Y/N)
- Oral corticosteroid (Y/N)

- Corticosteroid injection (Y/N)
- Physical therapy (Y/N)
- Chiropractic visit (Y/N)
- Outpatient encounter with OA diagnosis code of any joint (Y/N)
- Number of outpatient encounters with an M diagnosis code

### Appendix E. HCRU types and definitions

We measured HCRU by interactions with the healthcare system (cumulative count data where appropriate). HCRU was captured longitudinally:

- From
  - The day after the Procedure END date
  - 90 days after the Procedure START date
    - Only to be used for bundled payment strategy when calculating costs for TKA/pTKA
- Until
  - 12 months after the Procedure START date
  - 24 months after the Procedure START date
  - 36 months after the Procedure START date
  - 48 months after the Procedure START date

The START and END dates of an index Procedure were defined as the START and END dates of that procedure's episode of care. Episodes of care were defined as follows:

- For outpatient claims, the episode of care starts and ends on the first service date of the claim
- For inpatient claims (including ER claims), claims that are not separated by at least one full day will count as part of the same episode of care. For example, see the table below

| Patient | Claim place of service | Claim first service date | Claim last service date | Episode of care |
|---|---|---|---|---|
| A | ER | 1/1/2019 | 1/1/2019 | 1 |
| A | Inpatient | 1/2/2019 | 1/5/2019 | 1 |
| A | Inpatient | 1/4/2019 | 1/7/2019 | 1 |
| A | Inpatient | 1/9/2019 | 1/10/2019 | 2 |
| A | ER | 1/11/2019 | 1/12/2019 | 2 |
| A | Inpatient | 1/13/2019 | 1/14/2019 | 2 |

Some index procedures had both inpatient and outpatient claims. For example, a patient may have an outpatient claim with CPT code 29880 (meniscectomy) and an inpatient claim with CPT code 29881 (also meniscectomy) on the same day. In these cases, the procedure was counted as inpatient.

Encounter types of interest are listed below. The number of encounters were counted as the number of unique episodes of care, except for radiology variables and subsequent BMAC and PRP procedures. For radiology variables, the number of encounters were counted as the number of unique claim First Service Dates. For subsequent BMAC and PRP procedures, the number of procedures were counted as the number of unique Primary Procedure Dates. Note that, for outpatient claims, counting unique episodes of care is the same thing as counting unique claim First Service Dates.

- Outpatient (non-physical therapy) visits with a relevant orthopedic diagnosis code
  - Relevant codes are the M and S ICD-10-CM codes listed Appendix D plus after care codes: Z96.651, Z96.652, Z47.1, Z47.89, Z48.89
  - Outpatient visits were considered non-physical therapy so long as a physical therapy outpatient visit did not occur on the same day
- Outpatient physical therapy with a relevant orthopedic diagnosis code
  - Relevant codes are the M and S ICD-10-CM codes listed in Appendix D plus after care codes: Z96.651, Z96.652, Z47.1, Z47.89, Z48.89
- Radiology regardless of contrast, i.e. with contrast and without contrast were grouped together
  - Xray of knee joint
  - MRI of lower joint
  - CT of lower extremity
- Subsequent surgical knee procedures on the same knee or unknown side. Inclusive of codes for revision and draining. Outpatient procedures that occur during an inpatient episode of care will be counted as part of that episode of care for the purposes of counting unique encounters
  - Subsequent TKA/pTKA (denovo procedure or revision)
  - Subsequent Meniscectomy
- Subsequent PRP and BMAC procedures on the same knee or unknown side.
- Emergency Department or Inpatient care for:
  - Deep vein thrombosis, pulmonary embolism, or other post-surgical embolism
  - Hematoma
  - Infection
- Opioid medications
  - Number of unique prescription fill dates

### Appendix F. Costing

**Medicare cost methodology.** Costs are aggregated beginning at the procedure date and summed over time within the discrete time frame of interest. Costs will be assigned to each type of HCRU based on estimates of Medicare cost from external references and literature (Table 1F). The Medicare cost of TKA/pTKA was generally reported as a 90-day bundled payment and therefore, no other costs were not counted until AFTER 90 days (that is no HCRU accrued a cost until after 90 days if TKA/pTKA was the initial procedure costed at $25,000.

**Table 1F. Medicare cost assignment based on external literature.**

| Service | Cost assigned | References |
|---|---|---|
| TKA/pTKA | $25,000 (DRG bundled payment) | $17K-$29K with mean $22K in external references from 2013,[1] and total joint replacement $25K in 2020.[2] Assigned $25,000 allowing for inflation. |
| Arthroscopic Meniscectomy | $3,323 | *Weighted average of $3900 and $2250 at 65% and 35% respectively HOPD and ASC.[3] |
| Knee MRI without contrast | $250 | $191-$302[4] |
| Knee CT with/out contrast | $200 | $150-$170, $209-$275[5] |
| Knee x-ray | $50 | $35-46 (outpatient)[6] |
| Outpatient visit | $100 | $50-$360[7] based on Kaiser estimated fees (Medicare not reported) |
| Physical Therapy | $100 | $50-170[7], $30-$100[8] |
| Hospitalization for DVT, PE, or other embolism | $10,000 | $10,000-$13,500 average for managed care payers in 2007 (Medicare not reported)[9] |
| Hospitalization for hematoma | $19,000 | Not directly reported. Estimated as difference in Medicare payment for TKA with and without complication (17-22K)[11] |
| Hospitalization for post-procedural infection | $25,000 | $20,000 in 2018 plus approximately 25% for subsequent SNF care[10] |
| Regenexx procedure | PRP: $3085.47<br>BMAC: $11,984.61 | |

1. Ellimoottil C, Ryan AM, Hou H, Dupree JM, Hallstrom B, Miller DC. Implications of the Definition of an Episode of Care Used in the Comprehensive Care for Joint Replacement Model. JAMA Surg. 2017 Jan 1;152(1):49-54. doi: 10.1001/jamasurg.2016.3098. PMID: 27682525; PMCID: PMC5336141.
2. Chen DQ, Parvataneni HK, Miley EN, Deen JT, Pulido LF, Prieto HA, Gray CF. Lessons Learned From the Comprehensive Care for Joint Replacement Model at an Academic Tertiary Center: The Good, the Bad, and the Ugly. J Arthroplasty. 2023 Jul;38(7 Suppl 2):S54-S62. doi: 10.1016/j.arth.2023.02.014. Epub 2023 Feb 11. PMID: 36781061; PMCID: PMC10839807.
3. https://www.medicare.gov/procedure-price-lookup/cost/29882/#:~:text=29882

**Multiplier methodology.** Medicare based costs were multiplied by 2 (which is in the range of multipliers across different cost settings).

**Table 2F. Multipliers used for private payer costing estimate**

| Service Type | Commercial:Medicare Payment Ratio | Source (Typical) |
|---|---|---|
| Inpatient hospital | 1.8 to 2.5 | RAND Hospital Price Transparency Studies |
| Outpatient hospital | 2.0 to 3.0 | MedPAC, RAND, HCCI |
| Physician services | 1.2 to 1.8 | MedPAC, CMS Office of the Actuary |
| Ambulatory surgery | 1.4 to 2.0 | HCCI, FAIR Health |

**Private payer average cost methodology.** Using COMMERCIAL PAYER data, we obtained the average cost for an episode of care associated with each type of healthcare resource. For example, a surgical procedure might have physician billing and facility billing separately, along with a different bill for anesthesia as for the surgeon. Multiple claims for the same episode of care will be treated as a single health care use (and counted towards more expensive/acute visit type) and all applied costs will be summed to compute the total cost of care. Table 3F shows the average cost of care for each type of service among patients receiving orthopedic care in the COMMERCIAL PAYER data.

**Table 3F. Commercial payer cost assignment; based average allowed amounts in commercial data.**

| Service | Cost assigned |
|---|---|
| TKA/pTKA | $22,324 |
| Arthroscopic Meniscectomy | $5955 |
| Outpatient visit | $392 |
| Physical Therapy | $106 |
| Knee x-ray | $87 |
| Knee CT without contrast | $454 |
| Knee MRI without contrast | $667 |
| Hospitalization for DVT, PE, or other embolism | $25,693 |
| Hospitalization for hematoma | $31,907 |
| Hospitalization for post-procedural infection | $34,395 |
| Regenexx procedure | 2023 PRP: $3085.47<br>2023 BMAC: $11,984.61 |

**Computation of aggregate costs.**

Each occurrence of a relevant HCRU was assigned a cost and all costs were aggregated over the discrete time frame of interest (1,2,3,4 years). The cost of the initial procedure was also estimated and included in the sum. When calculating Medicare costs, reimbursement for inpatient procedures is typically based on a bundled payment program (lump sum that covers all related costs for 90 days) rather than fee for service (or individual service costs). This impacted the published estimates of costs for TKA/pKA which is frequently an inpatient service. The literature on TKA/pKA reports a bundled payment. Therefore, we used that bundled payment amount for the cost of TKA/pKA (in the Medicare framework) and did not accrue any additional costs until after 90 days, when costing TKA/pKA. Overlapping records of identical visit types were treated as a single health care use (and counted towards more expensive/acute visit type) and all applied costs were summed; e.g. two outpatient visits on same day counted once towards the tally of outpatient visit HCRU type, and the associated cost of this single tally was the sum of any costs at either visit. A lack of utilization data was treated as zero (0) utilization and thus attributed $0 in costs.

**Appendix H. Pre-matching demographic and clinical characteristics – BMAC with or without PRP versus TKA/pKA**

| **Characteristic** | **BMAC ± PRP**<br>N = 185 | **TKA/PKA**<br>N = 97,327 | **Overall**<br>N = 97,512 |
|---|---|---|---|
| Continuous commercial health plan enrollment after index procedure | | | |
| CE for 12 months | 185 (100.0%) | 97,327 (100.0%) | 97,512 (100.0%) |
| CE for 24 months | 130 (70.3%) | 63,160 (64.9%) | 63,290 (64.9%) |
| CE for 36 months | 94 (50.8%) | 40,091 (41.2%) | 40,185 (41.2%) |
| CE for 48 months | 70 (37.8%) | 24,198 (24.9%) | 24,268 (24.9%) |
| Location | | | |
| Right | 54 (29.2%) | 50,416 (51.8%) | 50,470 (51.8%) |
| Left | 67 (36.2%) | 46,911 (48.2%) | 46,978 (48.2%) |
| Both | 64 (34.6%) | -- | 64 (0.1%) |
| Sex | | | |
| Female | 83 (44.9%) | 53,840 (55.3%) | 53,923 (55.3%) |
| Male | 102 (55.1%) | 43,485 (44.7%) | 43,587 (44.7%) |
| Unknown | <11 | <11 | <11 |
| Age | | | |
| Mean (SD) | 56.1 (8.3) | 59.4 (6.2) | 59.3 (6.2) |
| Median (Q1, Q3) | 58.0 (53.0, 62.0) | 60.0 (56.0, 63.0) | 60.0 (56.0, 63.0) |
| Min, Max | 21.0, 75.0 | 19.0, 84.0 | 19.0, 84.0 |
| Geographic region | | | |
| Northeast | 34 (18.4%) | 21,690 (22.3%) | 21,724 (22.3%) |
| Midwest | 89 (48.1%) | 44,010 (45.2%) | 44,099 (45.2%) |
| South | 22 (11.9%) | 24,250 (24.9%) | 24,272 (24.9%) |
| West | 40 (21.6%) | 7,299 (7.5%) | 7,339 (7.5%) |
| Other | <11 | 78 (0.1%) | -- |
| Injury to the treated knee within 12 months prior to the procedure | | | |
| Meniscus tear | 57 (30.8%) | 8,909 (9.2%) | 8,966 (9.2%) |
| Sprain of collateral ligament of knee | 11 (5.9%) | 1,109 (1.1%) | 1,120 (1.1%) |
| Sprain of cruciate ligament of knee | 13 (7.0%) | 1,177 (1.2%) | 1,190 (1.2%) |
| Other knee injuries (cartilage tear, subluxations/dislocations, sprains) | <11 | 1,609 (1.7%) | -- |
| Orthopedic diagnoses on the treated knee within 12 months prior to the procedure | | | |
| Osteoarthritis | 185 (100.0%) | 97,327 (100.0%) | 97,512 (100.0%) |
| Acquired deformity of the knee | <11 | 4,674 (4.8%) | -- |
| Chondromalacia patellae | 32 (17.3%) | 4,791 (4.9%) | 4,823 (4.9%) |
| Other disorders of patella | <11 | 140 (0.1%) | -- |
| Derangement of meniscus | 43 (23.2%) | 4,484 (4.6%) | 4,527 (4.6%) |
| Derangement, instability | 23 (12.4%) | 1,600 (1.6%) | 1,623 (1.7%) |
| Spontaneous disruption of ligaments | <11 | 241 (0.2%) | -- |
| Severe derangement | <11 | 1,104 (1.1%) | -- |
| Derangement, unspecified | <11 | 1,982 (2.0%) | -- |
| Musculoskeletal conditions (not exclusive to the treated knee) within 12 months prior to the procedure | | | |
| Knee effusion | 66 (35.7%) | 16,887 (17.4%) | 16,953 (17.4%) |
| Number of knee effusions | | | |
| Mean (SD) | 0.6 (1.7) | 0.3 (1.1) | 0.3 (1.1) |
| Median (Q1, Q3) | 0.0 (0.0, 1.0) | 0.0 (0.0, 0.0) | 0.0 (0.0, 0.0) |
| Min, Max | 0.0, 19.0 | 0.0, 80.0 | 0.0, 80.0 |
| Number of knee effusions among patients who had at least one | | | |
| Mean (SD) | 1.8 (2.4) | 1.7 (2.1) | 1.7 (2.1) |
| Median (Q1, Q3) | 1.0 (1.0, 2.0) | 1.0 (1.0, 2.0) | 1.0 (1.0, 2.0) |
| Min, Max | 1.0, 19.0 | 1.0, 80.0 | 1.0, 80.0 |
| Missing | 119 | 80,440 | 80,559 |
| Enthesopathy of the lower limb, excluding foot | 22 (11.9%) | 3,749 (3.9%) | 3,771 (3.9%) |

| Characteristic | BMAC ± PRP<br>N = 185 | TKA/PKA<br>N = 97,327 | Overall<br>N = 97,512 |
|---|---|---|---|
| Other knee joint disorders (pain in knee, bursopathies, enthesopathies, stiffness, other instability, osteophytes) | 139 (75.1%) | 73,550 (75.6%) | 73,689 (75.6%) |
| Disorders of synovium and tendon | 11 (5.9%) | 1,349 (1.4%) | 1,360 (1.4%) |
| Other soft tissue disorders | 54 (29.2%) | 31,796 (32.7%) | 31,850 (32.7%) |
| Biomechanical lesions | <11 | 634 (0.7%) | -- |
| Dorsopathy | 77 (41.6%) | 31,588 (32.5%) | 31,665 (32.5%) |
| Osteoarthritis elsewhere | 31 (16.8%) | 25,534 (26.2%) | 25,565 (26.2%) |
| Injury to other parts of the body | 35 (18.9%) | 15,953 (16.4%) | 15,988 (16.4%) |
| Injury to adjacent parts of the body | 12 (6.5%) | 5,651 (5.8%) | 5,663 (5.8%) |
| ACL/MCL indication | 25 (13.5%) | 2,324 (2.4%) | 2,349 (2.4%) |
| General comorbidities documented in the 12 months prior to the procedure | | | |
| Obesity | 32 (17.3%) | 38,972 (40.0%) | 39,004 (40.0%) |
| Diabetes | <11 | 18,714 (19.2%) | -- |
| Depression/Anxiety/Bipolar | 29 (15.7%) | 20,805 (21.4%) | 20,834 (21.4%) |
| Medications within 12 months prior to the procedure | | | |
| Opioid medication | 57 (30.8%) | 28,680 (29.5%) | 28,737 (29.5%) |
| NSAID medication | 43 (23.2%) | 35,853 (36.8%) | 35,896 (36.8%) |
| Tramadol medication | <11 | 11,092 (11.4%) | -- |
| Neuropathic pain medication | 16 (8.6%) | 13,873 (14.3%) | 13,889 (14.2%) |
| Interventions in the 12 months prior to the procedure | | | |
| Oral corticosteroids | 31 (16.8%) | 20,562 (21.1%) | 20,593 (21.1%) |
| Corticosteroid injection | 67 (36.2%) | 57,765 (59.4%) | 57,832 (59.3%) |
| Number of corticosteroid injections | | | |
| Mean (SD) | 0.6 (1.0) | 1.2 (1.5) | 1.2 (1.5) |
| Median (Q1, Q3) | 0.0 (0.0, 1.0) | 1.0 (0.0, 2.0) | 1.0 (0.0, 2.0) |
| Min, Max | 0.0, 8.0 | 0.0, 28.0 | 0.0, 28.0 |
| Hyaluronic acid injection | 26 (14.1%) | 15,412 (15.8%) | 15,438 (15.8%) |
| Number of hyaluronic acid injections | | | |
| Mean (SD) | 0.3 (0.8) | 0.4 (1.0) | 0.4 (1.0) |
| Median (Q1, Q3) | 0.0 (0.0, 0.0) | 0.0 (0.0, 0.0) | 0.0 (0.0, 0.0) |
| Min, Max | 0.0, 5.0 | 0.0, 18.0 | 0.0, 18.0 |
| Physical therapy | 87 (47.0%) | 36,603 (37.6%) | 36,690 (37.6%) |
| Number of PT visits | | | |
| Mean (SD) | 4.9 (9.6) | 2.7 (6.9) | 2.7 (7.0) |
| Median (Q1, Q3) | 0.0 (0.0, 6.0) | 0.0 (0.0, 1.0) | 0.0 (0.0, 1.0) |
| Min, Max | 0.0, 63.0 | 0.0, 150.0 | 0.0, 150.0 |
| Health seeking behavior in the 12 months prior to the procedure | | | |
| Outpatient visit | 185 (100.0%) | 97,327 (100.0%) | 97,512 (100.0%) |
| Number of outpatient visits | | | |
| Mean (SD) | 20.4 (16.1) | 20.4 (14.5) | 20.4 (14.5) |
| Median (Q1, Q3) | 15.0 (9.0, 26.0) | 17.0 (11.0, 26.0) | 17.0 (11.0, 26.0) |
| Min, Max | 1.0, 90.0 | 1.0, 360.0 | 1.0, 360.0 |
| ER visit | 23 (12.4%) | 11,753 (12.1%) | 11,776 (12.1%) |
| Number of ER visits | | | |
| Mean (SD) | 0.1 (0.4) | 0.2 (0.8) | 0.2 (0.8) |
| Median (Q1, Q3) | 0.0 (0.0, 0.0) | 0.0 (0.0, 0.0) | 0.0 (0.0, 0.0) |
| Min, Max | 0.0, 2.0 | 0.0, 159.0 | 0.0, 159.0 |
| Chiropractic visit | 41 (22.2%) | 11,712 (12.0%) | 11,753 (12.1%) |
| Number of chiropractic visits | | | |
| Mean (SD) | 2.2 (5.6) | 1.1 (4.3) | 1.1 (4.3) |
| Median (Q1, Q3) | 0.0 (0.0, 0.0) | 0.0 (0.0, 0.0) | 0.0 (0.0, 0.0) |
| Min, Max | 0.0, 33.0 | 0.0, 118.0 | 0.0, 118.0 |
| Knee x-ray | 128 (69.2%) | 90,933 (93.4%) | 91,061 (93.4%) |
| MRI of lower joint | 112 (60.5%) | 19,364 (19.9%) | 19,476 (20.0%) |

| Characteristic | BMAC ± PRP<br>N = 185 | TKA/PKA<br>N = 97,327 | Overall<br>N = 97,512 |
|---|---|---|---|
| Knee x-ray or MRI of lower joint | 163 (88.1%) | 91,965 (94.5%) | 92,128 (94.5%) |
| Other severity proxies in the 12 months prior to the procedure | | | |
| Osteoarthritis outpatient visit for the treated knee | 185 (100.0%) | 97,319 (100.0%) | 97,504 (100.0%) |
| Number of osteoarthritis outpatient visits | | | |
| Mean (SD) | 3.9 (3.3) | 5.5 (4.2) | 5.5 (4.2) |
| Median (Q1, Q3) | 3.0 (2.0, 5.0) | 4.0 (3.0, 7.0) | 4.0 (3.0, 7.0) |
| Min, Max | 1.0, 25.0 | 0.0, 115.0 | 0.0, 115.0 |
| Number of OA outpatient visits for the treated knee | | | |
| Mean (SD) | 3.5 (2.7) | 4.8 (3.6) | 4.8 (3.6) |
| Median (Q1, Q3) | 3.0 (2.0, 4.0) | 4.0 (3.0, 6.0) | 4.0 (3.0, 6.0) |
| Min, Max | 1.0, 14.0 | 0.0, 91.0 | 0.0, 91.0 |
| Outpatient visit with an 'M' dx code | 185 (100.0%) | 97,326 (100.0%) | 97,511 (100.0%) |
| Number of outpatient visits with an 'M' dx code | | | |
| Mean (SD) | 11.3 (11.3) | 10.1 (9.3) | 10.1 (9.3) |
| Median (Q1, Q3) | 8.0 (4.0, 14.0) | 7.0 (4.0, 12.0) | 7.0 (4.0, 12.0) |
| Min, Max | 1.0, 68.0 | 0.0, 153.0 | 0.0, 153.0 |

**Appendix I. Pre-matching demographic and clinical characteristics – PRP versus Meniscectomy**

| **Characteristic** | **PRP**<br>N = 167 | **Meniscectomy**<br>N = 116,473 | **Overall**<br>N = 116,640 |
|---|---|---|---|
| Continuous commercial health plan enrollment after index procedure | | | |
| CE for 12 months | 167 (100.0%) | 116,473 (100.0%) | 116,640 (100.0%) |
| CE for 24 months | 105 (62.9%) | 84,870 (72.9%) | 84,975 (72.9%) |
| CE for 36 months | 63 (37.7%) | 60,342 (51.8%) | 60,405 (51.8%) |
| CE for 48 months | 35 (21.0%) | 40,936 (35.1%) | 40,971 (35.1%) |
| Location | | | |
| Right | 60 (35.9%) | 59,822 (51.4%) | 59,882 (51.3%) |
| Left | 63 (37.7%) | 56,651 (48.6%) | 56,714 (48.6%) |
| Both | 44 (26.3%) | -- | 44 (0.0%) |
| Sex | | | |
| Female | 80 (47.9%) | 49,197 (42.2%) | 49,277 (42.2%) |
| Male | 87 (52.1%) | 67,276 (57.8%) | 67,363 (57.8%) |
| Age | | | |
| Mean (SD) | 51.6 (11.0) | 49.9 (11.4) | 49.9 (11.4) |
| Median (Q1, Q3) | 54.0 (46.0, 59.0) | 52.0 (44.0, 58.0) | 52.0 (44.0, 58.0) |
| Min, Max | 18.0, 72.0 | 18.0, 84.0 | 18.0, 84.0 |
| Geographic region | | | |
| Northeast | 39 (23.4%) | 25,394 (21.8%) | 25,433 (21.8%) |
| Midwest | 69 (41.3%) | 49,827 (42.8%) | 49,896 (42.8%) |
| South | 37 (22.2%) | 29,294 (25.2%) | 29,331 (25.1%) |
| West | 22 (13.2%) | 11,861 (10.2%) | 11,883 (10.2%) |
| Other | <11 | 97 (0.1%) | -- |
| Injury to the treated knee within 12 months prior to the procedure | | | |
| Meniscus tear | 23 (13.8%) | 93,708 (80.5%) | 93,731 (80.4%) |
| Sprain of collateral ligament of knee | <11 | <11 | -- |
| Sprain of cruciate ligament of knee | <11 | <11 | -- |
| Other knee injuries (cartilage tear, subluxations/dislocations, sprains) | <11 | 8,827 (7.6%) | -- |
| Orthopedic diagnoses on the treated knee within 12 months prior to the procedure | | | |
| Osteoarthritis | 48 (28.7%) | 29,179 (25.1%) | 29,227 (25.1%) |
| Acquired deformity of the knee | <11 | 231 (0.2%) | -- |
| Chondromalacia patellae | 30 (18.0%) | 24,392 (20.9%) | 24,422 (20.9%) |
| Other disorders of patella | <11 | 673 (0.6%) | -- |
| Derangement of meniscus | 14 (8.4%) | 32,073 (27.5%) | 32,087 (27.5%) |
| Derangement, instability | <11 | 5,329 (4.6%) | -- |
| Spontaneous disruption of ligaments | <11 | <11 | -- |
| Severe derangement | <11 | 4,274 (3.7%) | -- |
| Derangement, unspecified | <11 | 13,014 (11.2%) | -- |
| Musculoskeletal conditions (not exclusive to the treated knee) within 12 months prior to the procedure | | | |
| Knee effusion | 23 (13.8%) | 37,092 (31.8%) | 37,115 (31.8%) |
| Number of knee effusions | | | |
| Mean (SD) | 0.2 (0.6) | 0.5 (1.0) | 0.5 (1.0) |
| Median (Q1, Q3) | 0.0 (0.0, 0.0) | 0.0 (0.0, 1.0) | 0.0 (0.0, 1.0) |
| Min, Max | 0.0, 3.0 | 0.0, 58.0 | 0.0, 58.0 |
| Number of knee effusions among patients who had at least one | | | |
| Mean (SD) | 1.5 (0.6) | 1.5 (1.3) | 1.5 (1.3) |
| Median (Q1, Q3) | 1.0 (1.0, 2.0) | 1.0 (1.0, 2.0) | 1.0 (1.0, 2.0) |
| Min, Max | 1.0, 3.0 | 1.0, 58.0 | 1.0, 58.0 |
| Missing | 144 | 79,381 | 79,525 |
| Enthesopathy of the lower limb, excluding foot | 21 (12.6%) | 6,186 (5.3%) | 6,207 (5.3%) |

| Characteristic | PRP<br>N = 167 | Meniscectomy<br>N = 116,473 | Overall<br>N = 116,640 |
|---|---|---|---|
| Other knee joint disorders (pain in knee, bursopathies, enthesopathies, stiffness, other instability, osteophytes) | 103 (61.7%) | 101,612 (87.2%) | 101,715 (87.2%) |
| Disorders of synovium and tendon | <11 | 4,942 (4.2%) | -- |
| Other soft tissue disorders | 76 (45.5%) | 35,001 (30.1%) | 35,077 (30.1%) |
| Biomechanical lesions | <11 | 943 (0.8%) | -- |
| Dorsopathy | 76 (45.5%) | 32,363 (27.8%) | 32,439 (27.8%) |
| Osteoarthritis elsewhere | 34 (20.4%) | 10,175 (8.7%) | 10,209 (8.8%) |
| Injury to other parts of the body | 46 (27.5%) | 21,879 (18.8%) | 21,925 (18.8%) |
| Injury to adjacent parts of the body | 17 (10.2%) | 10,044 (8.6%) | 10,061 (8.6%) |
| ACL/MCL indication | <11 | <11 | -- |
| General comorbidities documented in the 12 months prior to the procedure | | | |
| Obesity | 22 (13.2%) | 26,944 (23.1%) | 26,966 (23.1%) |
| Diabetes | 16 (9.6%) | 10,859 (9.3%) | 10,875 (9.3%) |
| Depression/Anxiety/Bipolar | 26 (15.6%) | 20,911 (18.0%) | 20,937 (18.0%) |
| Medications within 12 months prior to the procedure | | | |
| Opioid medication | 32 (19.2%) | 31,529 (27.1%) | 31,561 (27.1%) |
| NSAID medication | 31 (18.6%) | 32,327 (27.8%) | 32,358 (27.7%) |
| Tramadol medication | <11 | 7,328 (6.3%) | -- |
| Neuropathic pain medication | 16 (9.6%) | 8,493 (7.3%) | 8,509 (7.3%) |
| Interventions in the 12 months prior to the procedure | | | |
| Oral corticosteroids | 26 (15.6%) | 23,931 (20.5%) | 23,957 (20.5%) |
| Corticosteroid injection | 41 (24.6%) | 42,080 (36.1%) | 42,121 (36.1%) |
| Number of corticosteroid injections | | | |
| Mean (SD) | 0.4 (0.8) | 0.6 (1.0) | 0.6 (1.0) |
| Median (Q1, Q3) | 0.0 (0.0, 0.0) | 0.0 (0.0, 1.0) | 0.0 (0.0, 1.0) |
| Min, Max | 0.0, 4.0 | 0.0, 71.0 | 0.0, 71.0 |
| Hyaluronic acid injection | <11 | 592 (0.5%) | -- |
| Number of hyaluronic acid injections | | | |
| Mean (SD) | 0.0 (0.2) | 0.0 (0.2) | 0.0 (0.2) |
| Median (Q1, Q3) | 0.0 (0.0, 0.0) | 0.0 (0.0, 0.0) | 0.0 (0.0, 0.0) |
| Min, Max | 0.0, 3.0 | 0.0, 10.0 | 0.0, 10.0 |
| Physical therapy | 70 (41.9%) | 31,941 (27.4%) | 32,011 (27.4%) |
| Number of PT visits | | | |
| Mean (SD) | 5.6 (14.3) | 2.3 (6.4) | 2.3 (6.4) |
| Median (Q1, Q3) | 0.0 (0.0, 6.0) | 0.0 (0.0, 1.0) | 0.0 (0.0, 1.0) |
| Min, Max | 0.0, 153.0 | 0.0, 140.0 | 0.0, 153.0 |
| Health seeking behavior in the 12 months prior to the procedure | | | |
| Outpatient visit | 166 (99.4%) | 116,328 (99.9%) | 116,494 (99.9%) |
| Number of outpatient visits | | | |
| Mean (SD) | 18.7 (17.3) | 16.0 (13.0) | 16.1 (13.1) |
| Median (Q1, Q3) | 14.0 (8.0, 24.0) | 12.0 (8.0, 20.0) | 12.0 (8.0, 20.0) |
| Min, Max | 0.0, 162.0 | 0.0, 364.0 | 0.0, 364.0 |
| ER visit | 18 (10.8%) | 14,615 (12.5%) | 14,633 (12.5%) |
| Number of ER visits | | | |
| Mean (SD) | 0.1 (0.4) | 0.2 (0.6) | 0.2 (0.6) |
| Median (Q1, Q3) | 0.0 (0.0, 0.0) | 0.0 (0.0, 0.0) | 0.0 (0.0, 0.0) |
| Min, Max | 0.0, 2.0 | 0.0, 29.0 | 0.0, 29.0 |
| Chiropractic visit | 37 (22.2%) | 15,605 (13.4%) | 15,642 (13.4%) |
| Number of chiropractic visits | | | |
| Mean (SD) | 3.1 (13.2) | 1.1 (4.2) | 1.1 (4.2) |
| Median (Q1, Q3) | 0.0 (0.0, 0.0) | 0.0 (0.0, 0.0) | 0.0 (0.0, 0.0) |
| Min, Max | 0.0, 153.0 | 0.0, 139.0 | 0.0, 153.0 |
| Knee x-ray | 59 (35.3%) | 102,751 (88.2%) | 102,810 (88.1%) |
| MRI of lower joint | 54 (32.3%) | 103,558 (88.9%) | 103,612 (88.8%) |

| Characteristic | PRP<br>N = 167 | Meniscectomy<br>N = 116,473 | Overall<br>N = 116,640 |
|---|---|---|---|
| Knee x-ray or MRI of lower joint | 79 (47.3%) | 112,042 (96.2%) | 112,121 (96.1%) |
| Other severity proxies in the 12 months prior to the procedure | | | |
| Osteoarthritis outpatient visit | 73 (43.7%) | 37,752 (32.4%) | 37,825 (32.4%) |
| Number of osteoarthritis outpatient visits | | | |
| Mean (SD) | 0.8 (1.6) | 0.5 (1.2) | 0.5 (1.2) |
| Median (Q1, Q3) | 0.0 (0.0, 1.0) | 0.0 (0.0, 1.0) | 0.0 (0.0, 1.0) |
| Min, Max | 0.0, 9.0 | 0.0, 80.0 | 0.0, 80.0 |
| Number of OA outpatient visits for the treated knee | 48 (28.7%) | 29,134 (25.0%) | 29,182 (25.0%) |
| Outpatient visit with an 'M' dx code | 160 (95.8%) | 113,566 (97.5%) | 113,726 (97.5%) |
| Number of outpatient visits with an 'M' dx code | | | |
| Mean (SD) | 9.4 (10.6) | 6.9 (8.1) | 6.9 (8.1) |
| Median (Q1, Q3) | 5.0 (2.0, 12.0) | 4.0 (3.0, 8.0) | 4.0 (3.0, 8.0) |
| Min, Max | 0.0, 55.0 | 0.0, 178.0 | 0.0, 178.0 |

**Appendix J. Healthcare Resource Utilization 36 months post procedure – BMAC with or without PRP versus TKA/pKA**

| **Characteristic** | **BMAC ± PRP**<br>N = 80 | **TKA/PKA**<br>N = 734 | **Overall**<br>N = 814 | **p-value** |
|---|---|---|---|---|
| Number of outpatient visits | | | | 0.03 |
| Mean (SD) | 11.6 (10.9) | 14.3 (14.2) | 14.0 (14.0) | |
| Median (Q1, Q3) | 8.0 (4.0, 16.0) | 10.0 (5.0, 18.0) | 10.0 (5.0, 18.0) | |
| Min, Max | 0.0, 50.0 | 0.0, 161.0 | 0.0, 161.0 | |
| Number of physical therapy visits | | | | 0.03 |
| Mean (SD) | 17.7 (21.2) | 20.1 (20.9) | 19.8 (20.9) | |
| Median (Q1, Q3) | 9.0 (0.0, 25.5) | 15.0 (4.0, 28.0) | 15.0 (4.0, 28.0) | |
| Min, Max | 0.0, 83.0 | 0.0, 184.0 | 0.0, 184.0 | |
| Knee x-ray | 32 (40.0%) | 704 (95.9%) | 736 (90.4%) | < 0.001 |
| MRI of lower joint | 27 (33.8%) | 64 (8.7%) | 91 (11.2%) | < 0.001 |
| CT of lower extremity | <11 | 56 (7.6%) | -- | 0.53 |
| Subsequent same side TKA/PKA | <11 | 82 (11.2%) | -- | 0.64 |
| Subsequent same side Meniscectomy | <11 | <11 | -- | -- |
| Subsequent same side ACL/MCL procedure | <11 | <11 | -- | -- |
| Subsequent orthobiologic procedure - any knee | 13 (16.3%) | -- | -- | -- |
| Subsequent orthobiologic procedure – same knee | <11 | -- | -- | -- |
| Deep vein thrombosis inpatient care | <11 | <11 | -- | -- |
| Hematoma inpatient care | <11 | <11 | -- | -- |
| Infection inpatient care | <11 | 13 (1.8%) | -- | -- |
| Pulmonary embolism inpatient care | <11 | <11 | -- | -- |
| Other post-surgical embolism inpatient care | <11 | <11 | -- | -- |
| DVT, PE, or OE inpatient care | <11 | <11 | -- | -- |
| Opioid medication | 42 (52.5%) | 354 (48.2%) | 396 (48.6%) | 0.54 |

**Appendix K. Healthcare Resource Utilization 48 months post procedure – BMAC with or without PRP versus TKA/pKA**

| **Characteristic** | **BMAC ± PRP**<br>N = 59 | **TKA/PKA**<br>N = 436 | **Overall**<br>N = 495 | **p-value** |
|---|---|---|---|---|
| Number of outpatient visits | | | | > 0.99 |
| Mean (SD) | 17.5 (15.6) | 17.9 (19.2) | 17.9 (18.8) | |
| Median (Q1, Q3) | 13.0 (6.0, 25.0) | 13.0 (6.0, 22.5) | 13.0 (6.0, 23.0) | |
| Min, Max | 0.0, 66.0 | 0.0, 174.0 | 0.0, 174.0 | |
| Number of physical therapy visits | | | | 0.5 |
| Mean (SD) | 23.6 (26.5) | 22.5 (25.1) | 22.6 (25.2) | |
| Median (Q1, Q3) | 13.0 (0.0, 41.0) | 15.5 (5.0, 31.0) | 15.0 (5.0, 31.0) | |
| Min, Max | 0.0, 109.0 | 0.0, 228.0 | 0.0, 228.0 | |
| Knee x-ray | 33 (55.9%) | 420 (96.3%) | 453 (91.5%) | < 0.001 |
| MRI of lower joint | 23 (39.0%) | 58 (13.3%) | 81 (16.4%) | < 0.001 |
| CT of lower extremity | <11 | 40 (9.2%) | -- | > 0.99 |
| Subsequent same side TKA/PKA | <11 | 51 (11.7%) | -- | 0.61 |
| Subsequent same side Meniscectomy | <11 | <11 | -- | -- |
| Subsequent same side ACL/MCL procedure | <11 | <11 | -- | -- |
| Subsequent orthobiologic procedure - any knee | <11 | -- | -- | -- |
| Subsequent orthobiologic procedure – same knee | <11 | -- | -- | -- |
| Deep vein thrombosis inpatient care | <11 | <11 | -- | -- |
| Hematoma inpatient care | <11 | <11 | -- | -- |
| Infection inpatient care | <11 | 12 (2.8%) | -- | -- |
| Pulmonary embolism inpatient care | <11 | <11 | -- | -- |
| Other post-surgical embolism inpatient care | <11 | <11 | -- | -- |
| DVT, PE, or OE inpatient care | <11 | 11 (2.5%) | -- | -- |
| Opioid medication | 34 (57.6%) | 222 (50.9%) | 256 (51.7%) | 0.41 |

**Appendix L. Costs – BMAC with or without PRP versus TKA/pKA at 36- and 48-Months Post-Procedure**

| Characteristic | BMAC ± PRP<br>N = 80 | TKA/pKA<br>N = 734 | Estimated mean difference (95% CI) | p-value |
|---|---|---|---|---|
| **36 months post-procedure** | | | | |
| External Medicare costs ($) | | | | |
| Mean (SD) | 19,520 (11,440) | 30,591 (10,435) | 11,071 (8,434, 13,708) | <0.001 |
| Median (Q1, Q3) | 14,885 (13,010, 19,610) | 26,625 (25,500, 28,900) | | |
| External Medicare cost ×2 ($) | | | | |
| Mean (SD) | 25,921 (21,856) | 61,182 (20,871) | 35,261 (30,203, 40,319) | <0.001 |
| Median (Q1, Q3) | 17,635 (14,035, 27,235) | 53,250 (51,000, 57,800) | | |
| Commercial payer costs ($) | | | | |
| Mean (SD) | 23,448 (13,934) | 34,259 (13,643) | 10,810 (7,578, 14,043) | <0.001 |
| Median (Q1, Q3) | 17,268 (14,057, 27,841) | 29,354 (26,491, 35,046) | | |
| Aggregate costing method ($) | | | | |
| Mean (SD) | 33,306 (51,688) | 45,438 (30,189) | 12,132 (509, 23,755) | 0.041 |
| Median (Q1, Q3) | 16,763 (13,827, 34,994) | 37,587 (27,897, 53,873) | | |
| Aggregate costing method – truncated ($) | | | | |
| Mean (SD) | 28,545 (22,006) | 43,378 (21,646) | 14,833 (9,726, 19,940) | <0.001 |
| Median (Q1, Q3) | 16,763 (13,827, 34,994) | 37,587 (27,897, 53,873) | | |
| **48 months post-procedure** | | | | |
| | **BMAC ± PRP**<br>N = 59 | **TKA/pKA**<br>N = 436 | **Estimated mean difference (95% CI)** | **p-value** |
| External Medicare costs ($) | | | | |
| Mean (SD) | 20,179 (12,114) | 31,974 (11,820) | 11,795 (8,477, 15,113) | <0.001 |
| Median (Q1, Q3) | 15,535 (13,285, 19,885) | 27,150 (25,750, 30,400) | | |
| External Medicare cost ×2 ($) | | | | |
| Mean (SD) | 27,555 (23,232) | 63,948 (23,641) | 36,393 (29,999, 42,787) | <0.001 |
| Median (Q1, Q3) | 19,085 (14,585, 27,785) | 54,300 (51,500, 60,800) | | |
| Commercial payer costs ($) | | | | |
| Mean (SD) | 25,694 (15,156) | 37,054 (16,073) | 11,360 (7,167, 15,553) | <0.001 |
| Median (Q1, Q3) | 20,315 (15,545, 30,133) | 30,930 (27,189, 39,113) | | |
| Aggregate costing method ($) | | | | |
| Mean (SD) | 32,224 (22,943) | 51,048 (47,350) | 18,825 (11,413, 26,236) | <0.001 |
| Median (Q1, Q3) | 22,706 (14,116, 43,452) | 41,341 (30,719, 58,572) | | |
| Aggregate costing method – truncated ($) | | | | |
| Mean (SD) | 32,265 (22,640) | 47,084 (23,520) | 14,819 (8,572, 21,066) | <0.001 |
| Median (Q1, Q3) | 22,706 (14,116, 43,452) | 41,341 (30,719, 58,572) | | |

**Appendix M. Healthcare Resource Utilization 36 months post procedure – PRP versus Meniscectomy**

| Characteristic | PRP<br>N = 63 | Meniscectomy<br>N = 842 | Overall<br>N = 905 | p-value |
|---|---|---|---|---|
| Number of outpatient visits | | | | 0.57 |
| Mean (SD) | 10.4 (12.2) | 10.5 (14.5) | 10.5 (14.4) | |
| Median (Q1, Q3) | 7.0 (2.0, 14.0) | 6.0 (2.0, 13.0) | 6.0 (2.0, 13.0) | |
| Min, Max | 0.0, 61.0 | 0.0, 141.0 | 0.0, 141.0 | |
| Number of physical therapy visits | | | | 0.45 |
| Mean (SD) | 21.0 (67.9) | 12.1 (17.7) | 12.7 (24.7) | |
| Median (Q1, Q3) | 7.0 (0.0, 18.0) | 5.0 (0.0, 16.0) | 5.0 (0.0, 16.0) | |
| Min, Max | 0.0, 532.0 | 0.0, 115.0 | 0.0, 532.0 | |
| Knee x-ray | 19 (30.2%) | 298 (35.4%) | 317 (35.0%) | 0.48 |
| MRI of lower joint | 19 (30.2%) | 174 (20.7%) | 193 (21.3%) | 0.11 |
| CT of lower extremity | <11 | 18 (2.1%) | -- | -- |
| Subsequent same side TKA/PKA | <11 | 44 (5.2%) | -- | -- |
| Subsequent same side Meniscectomy | <11 | 61 (7.2%) | -- | -- |
| Subsequent same side ACL/MCL procedure | <11 | <11 | -- | -- |
| Subsequent orthobiologic procedure - any knee | <11 | -- | -- | -- |
| Subsequent orthobiologic procedure – same knee | <11 | -- | -- | -- |
| Deep vein thrombosis inpatient care | <11 | <11 | -- | -- |
| Hematoma inpatient care | <11 | <11 | -- | -- |
| Infection inpatient care | <11 | 11 (1.3%) | -- | -- |
| Pulmonary embolism inpatient care | <11 | <11 | -- | -- |
| Other post-surgical embolism inpatient care | <11 | <11 | -- | -- |
| DVT, PE, or OE inpatient care | <11 | <11 | -- | -- |
| Opioid medication | 23 (36.5%) | 211 (25.1%) | 234 (25.9%) | 0.06 |

**Appendix N. Healthcare Resource Utilization 48 months post procedure – PRP versus Meniscectomy**

| Characteristic | PRP<br>N = 35 | Meniscectomy<br>N = 550 | Overall<br>N = 585 | p-value |
|---|---|---|---|---|
| Number of outpatient visits | | | | 0.95 |
| Mean (SD) | 12.3 (14.6) | 13.3 (19.4) | 13.3 (19.1) | |
| Median (Q1, Q3) | 8.0 (2.0, 15.0) | 7.0 (3.0, 16.0) | 7.0 (3.0, 16.0) | |
| Min, Max | 0.0, 72.0 | 0.0, 191.0 | 0.0, 191.0 | |
| Number of physical therapy visits | | | | 0.56 |
| Mean (SD) | 15.0 (15.7) | 15.1 (21.5) | 15.1 (21.2) | |
| Median (Q1, Q3) | 12.0 (0.0, 26.0) | 7.0 (0.0, 20.0) | 7.0 (0.0, 20.0) | |
| Min, Max | 0.0, 47.0 | 0.0, 129.0 | 0.0, 129.0 | |
| Knee x-ray | 13 (37.1%) | 233 (42.4%) | 246 (42.1%) | 0.67 |
| MRI of lower joint | 12 (34.3%) | 148 (26.9%) | 160 (27.4%) | 0.45 |
| CT of lower extremity | <11 | 15 (2.7%) | -- | -- |
| Subsequent same side TKA/PKA | <11 | 33 (6.0%) | -- | -- |
| Subsequent same side Meniscectomy | <11 | 39 (7.1%) | -- | -- |
| Subsequent same side ACL/MCL procedure | <11 | <11 | -- | -- |
| Subsequent orthobiologic procedure - any knee | <11 | -- | -- | -- |
| Subsequent orthobiologic procedure – same knee | <11 | -- | -- | -- |
| Deep vein thrombosis inpatient care | <11 | <11 | -- | -- |
| Hematoma inpatient care | <11 | <11 | -- | -- |
| Infection inpatient care | <11 | <11 | -- | -- |
| Pulmonary embolism inpatient care | <11 | <11 | -- | -- |
| Other post-surgical embolism inpatient care | <11 | <11 | -- | -- |
| DVT, PE, or OE inpatient care | <11 | <11 | -- | -- |
| Opioid medication | 13 (37.1%) | 148 (26.9%) | 161 (27.5%) | 0.26 |

### Appendix O. Costs – PRP versus Meniscectomy at 36- and 48-Months Post-Procedure

| Characteristic | PRP<br>N = 63 | Meniscectomy<br>N = 842 | Estimated mean difference (95% CI) | p-value |
|---|---|---|---|---|
| **36 months post-procedure** | | | | |
| External Medicare costs ($) | | | | |
| Mean (SD) | 7,267 (7,476) | 7,729 (8,130) | 462 (-1,481, 2,404) | 0.641 |
| Median (Q1, Q3) | 5,085 (3,785, 8,335) | 5,023 (3,923, 7,473) | | |
| External Medicare cost ×2 ($) | | | | |
| Mean (SD) | 10,721 (14,480) | 15,458 (16,259) | 4738 (964, 8,511) | 0.014 |
| Median (Q1, Q3) | 7,085 (4,485, 13,031) | 10,046 (7,846, 14,946) | | |
| Commercial payer costs ($) | | | | |
| Mean (SD) | 10,753 (9,883) | 13,867 (10,923) | 3,114 (542, 5,685) | 0.018 |
| Median (Q1, Q3) | 7,557 (4,961, 12,638) | 9,787 (7,494, 15,378) | | |
| Aggregate costing method ($) | | | | |
| Mean (SD) | 11,529 (11,934) | 14,725 (16,631) | 3,196 (16, 6,376) | 0.049 |
| Median (Q1, Q3) | 7,273 (4,654, 11,448) | 9,126 (5,607, 15,774) | | |
| Aggregate costing method – truncated ($) | | | | |
| Mean (SD) | 11,036 (9,979) | 13,443 (11,373) | 2,407 (-197, 5,011) | 0.07 |
| Median (Q1, Q3) | 7,273 (4,654, 11,448) | 9,126 (5,607, 15,774) | | |
| **48 months post-procedure** | | | | |
| | **PRP**<br>N = 35 | **Meniscectomy**<br>N = 550 | **Estimated mean difference (95% CI)** | **p-value** |
| External Medicare costs ($) | | | | |
| Mean (SD) | 6,840 (3,689) | 8,791 (9,644) | 1,951 (469, 3,433) | 0.01 |
| Median (Q1, Q3) | 5,385 (3,985, 8,685) | 5,548 (4,123, 8,473) | | |
| External Medicare cost ×2 ($) | | | | |
| Mean (SD) | 9,900 (5,985) | 17,583 (19,288) | 7,683 (5,098, 10,267) | <0.001 |
| Median (Q1, Q3) | 7,685 (4,885, 14,285) | 11,096 (8,246, 16,946) | | |
| Commercial payer costs ($) | | | | |
| Mean (SD) | 10,867 (7,375) | 15,911 (13,653) | 5,044 (2,309, 7,779) | <0.001 |
| Median (Q1, Q3) | 7,697 (5,353, 15,780) | 11,069 (7,947, 16,978) | | |
| Aggregate costing method ($) | | | | |
| Mean (SD) | 14,376 (18,745) | 16,479 (19,566) | 2,103 (-4,421, 8,626) | 0.527 |
| Median (Q1, Q3) | 7,587 (4,523, 13,186) | 10,014 (6,192, 18,536) | | |
| Aggregate costing method – truncated ($) | | | | |
| Mean (SD) | 13,107 (13,625) | 15,138 (13,638) | 2,031 (-2,698, 6,761) | 0.399 |
| Median (Q1, Q3) | 7,587 (4,523, 13,186) | 10,014 (6,192, 18,536) | | |

### Appendix P. Sensitivity analysis cohort flowchart

| Inclusion/exclusion criteria | Orthobiologic Procedures | TKA/pKA Procedures | Meniscectomy Procedures |
|---|---|---|---|
| Starting population: Receiving eligible procedures | 3833 | 340226 | 452957 |
| Procedure occurred between 1/1/2016 and 12/31/2023 | 3553 | 323702 | 438312 |
| Exclude patients who have multiple DOBs or sexes in the claims data | 3553 | 323691 | 438301 |
| Patients age 18 or older at the time of the procedure | 3503 | 323649 | 425899 |
| 12 months of continuous enrollment with commercial insurer after the procedure | 1149 | 220313 | 314196 |
| At least one outpatient claim in the 12 months after the procedure | 1101 | 220293 | 314194 |
| **Procedure is elective and preceded by a relevant clinical diagnosis** | | | |
| If occurs in hospital, occurs on first day of service | 1101 | 216837 | 314025 |
| **Exclude patients with clinical contra-indications** | | | |
| Patients with inflammatory syndrome diagnosis in the 12 months prior to the procedure | 1027 | 184541 | 284745 |
| Ineligible connective tissue disorder diagnosis in the 12 months prior to the procedure | 1014 | 181602 | 279923 |
| Ineligible joint disorder diagnosis in the 12 months prior to the procedure | 1014 | 181235 | 279348 |
| Ineligible injuries in the 12 months prior to the procedure | 1009 | 180143 | 278130 |
| Infectious arthropathies in the 12 months prior to the procedure | 1006 | 179627 | 277713 |
| Bone cancer in the 12 months prior to the procedure | 1005 | 179548 | 277663 |
| **Exclude patients with prior treatment contra-indications** | | | |
| Regenexx patients who had corticosteroids in the 2 months prior to the procedure | 922 | 179548 | 277663 |
| Previous knee surgery in the 12 months prior to the procedure | 898 | 144113 | 256365 |
| Mab medication in the 3 months prior to the procedure | 891 | 142456 | 254028 |
| Surgery patients who had Regenexx at any point | 891 | 142363 | 253860 |
| If a patient had multiple procedures, take the first one | 763 | 130466 | 241731 |

### Appendix Q. Orthobiologic procedures vs. TKA/PKA vs. Meniscectomy

| Characteristic | Orthobiologic procedures N = 763 | TKA/PKA N = 130,466 | Meniscectomy N = 241,731 | Overall N = 372,960 |
|---|---|---|---|---|
| Continuous commercial health plan enrollment after index procedure | | | | |
| CE for 12 months | 763 (100.0%) | 130,466 (100.0%) | 241,731 (100.0%) | 372,960 (100.0%) |
| CE for 24 months | 512 (67.1%) | 84,891 (65.1%) | 175,012 (72.4%) | 260,415 (69.8%) |
| CE for 36 months | 345 (45.2%) | 55,015 (42.2%) | 125,649 (52.0%) | 181,009 (48.5%) |
| CE for 48 months | 235 (30.8%) | 34,686 (26.6%) | 88,357 (36.6%) | 123,278 (33.1%) |
| Location | | | | |
| Right | 272 (35.6%) | 67,629 (51.8%) | 123,984 (51.3%) | 191,885 (51.4%) |
| Left | 282 (37.0%) | 62,837 (48.2%) | 117,747 (48.7%) | 180,866 (48.5%) |
| Both | 209 (27.4%) | -- | -- | 209 (0.1%) |
| Sex | | | | |
| Female | 340 (44.6%) | 72,721 (55.7%) | 108,224 (44.8%) | 181,285 (48.6%) |
| Male | 423 (55.4%) | 57,743 (44.3%) | 133,506 (55.2%) | 191,672 (51.4%) |
| Unknown | <11 | <11 | <11 | <11 |
| Age | | | | |
| Mean (SD) | 53.1 (10.7) | 59.1 (6.3) | 50.4 (11.2) | 53.5 (10.6) |
| Median (Q1, Q3) | 56.0 (49.0, 61.0) | 60.0 (56.0, 63.0) | 53.0 (45.0, 59.0) | 56.0 (49.0, 61.0) |
| Min, Max | 18.0, 78.0 | 18.0, 84.0 | 18.0, 84.0 | 18.0, 84.0 |
| Geographic region | | | | |
| Northeast | 163 (21.4%) | 27,669 (21.2%) | 54,723 (22.6%) | 82,555 (22.1%) |
| Midwest | 326 (42.7%) | 60,127 (46.1%) | 103,467 (42.8%) | 163,920 (44.0%) |
| South | 141 (18.5%) | 32,430 (24.9%) | 58,742 (24.3%) | 91,313 (24.5%) |
| West | 133 (17.4%) | 10,138 (7.8%) | 24,575 (10.2%) | 34,846 (9.3%) |
| Other | <11 | 102 (0.1%) | 224 (0.1%) | -- |
| Injury to the treated knee within 12 months prior to the procedure | | | | |
| Meniscus tear | 146 (19.1%) | 11,012 (8.4%) | 191,413 (79.2%) | 202,571 (54.3%) |
| Sprain of collateral ligament of knee | 24 (3.1%) | 1,347 (1.0%) | 18,359 (7.6%) | 19,730 (5.3%) |
| Sprain of cruciate ligament of knee | 42 (5.5%) | 1,483 (1.1%) | 11,386 (4.7%) | 12,911 (3.5%) |
| Other knee injuries (cartilage tear, subluxations/dislocations, sprains) | 20 (2.6%) | 1,997 (1.5%) | 18,783 (7.8%) | 20,800 (5.6%) |
| Orthopedic diagnoses on the treated knee within 12 months prior to the procedure | | | | |
| Osteoarthritis | 382 (50.1%) | 126,876 (97.2%) | 97,317 (40.3%) | 224,575 (60.2%) |
| Acquired deformity of the knee | <11 | 5,822 (4.5%) | 832 (0.3%) | -- |
| Chondromalacia patellae | 118 (15.5%) | 6,016 (4.6%) | 50,565 (20.9%) | 56,699 (15.2%) |
| Other disorders of patella | <11 | 176 (0.1%) | 1,383 (0.6%) | -- |
| Derangement of meniscus | 109 (14.3%) | 5,552 (4.3%) | 68,835 (28.5%) | 74,496 (20.0%) |
| Derangement, instability | 50 (6.6%) | 1,992 (1.5%) | 13,140 (5.4%) | 15,182 (4.1%) |
| Spontaneous disruption of ligaments | 16 (2.1%) | 310 (0.2%) | 1,899 (0.8%) | 2,225 (0.6%) |
| Severe derangement | <11 | 1,381 (1.1%) | 9,772 (4.0%) | -- |
| Derangement, unspecified | 20 (2.6%) | 2,513 (1.9%) | 26,516 (11.0%) | 29,049 (7.8%) |
| Musculoskeletal conditions (not exclusive to the treated knee) within 12 months prior to the procedure | | | | |
| Knee effusion | 160 (21.0%) | 20,978 (16.1%) | 80,940 (33.5%) | 102,078 (27.4%) |
| Number of knee effusions | | | | |
| Mean (SD) | 0.4 (1.2) | 0.3 (1.0) | 0.5 (1.2) | 0.4 (1.1) |
| Median (Q1, Q3) | 0.0 (0.0, 0.0) | 0.0 (0.0, 0.0) | 0.0 (0.0, 1.0) | 0.0 (0.0, 1.0) |
| Min, Max | 0.0, 19.0 | 0.0, 80.0 | 0.0, 134.0 | 0.0, 134.0 |
| Number of knee effusions among patients who had at least one | | | | |
| Mean (SD) | 1.7 (2.1) | 1.7 (2.1) | 1.5 (1.6) | 1.6 (1.7) |
| Median (Q1, Q3) | 1.0 (1.0, 2.0) | 1.0 (1.0, 2.0) | 1.0 (1.0, 2.0) | 1.0 (1.0, 2.0) |
| Min, Max | 1.0, 19.0 | 1.0, 80.0 | 1.0, 134.0 | 1.0, 134.0 |

| Characteristic | Orthobiologic procedures N = 763 | TKA/PKA N = 130,466 | Meniscectomy N = 241,731 | Overall N = 372,960 |
|---|---|---|---|---|
| Missing | 603 | 109,488 | 160,791 | 270,882 |
| Enthesopathy of the lower limb, excluding foot | 71 (9.3%) | 4,477 (3.4%) | 12,736 (5.3%) | 17,284 (4.6%) |
| Other knee joint disorders (pain in knee, bursopathies, enthesopathies, stiffness, other instability, osteophytes) | 437 (57.3%) | 94,300 (72.3%) | 205,581 (85.0%) | 300,318 (80.5%) |
| Disorders of synovium and tendon | 26 (3.4%) | 1,700 (1.3%) | 10,532 (4.4%) | 12,258 (3.3%) |
| Other soft tissue disorders | 218 (28.6%) | 38,992 (29.9%) | 70,497 (29.2%) | 109,707 (29.4%) |
| Biomechanical lesions | 11 (1.4%) | 761 (0.6%) | 1,810 (0.7%) | 2,582 (0.7%) |
| Dorsopathy | 264 (34.6%) | 39,458 (30.2%) | 64,495 (26.7%) | 104,217 (27.9%) |
| Osteoarthritis elsewhere | 111 (14.5%) | 32,577 (25.0%) | 23,154 (9.6%) | 55,842 (15.0%) |
| Injury to other parts of the body | 149 (19.5%) | 19,240 (14.7%) | 41,780 (17.3%) | 61,169 (16.4%) |
| Injury to adjacent parts of the body | 60 (7.9%) | 6,774 (5.2%) | 20,145 (8.3%) | 26,979 (7.2%) |
| ACL/MCL indication | 60 (7.9%) | 2,880 (2.2%) | 28,793 (11.9%) | 31,733 (8.5%) |
| General comorbidities documented in the 12 months prior to the procedure | | | | |
| Obesity | 89 (11.7%) | 48,230 (37.0%) | 56,071 (23.2%) | 104,390 (28.0%) |
| Diabetes | 61 (8.0%) | 23,932 (18.3%) | 23,599 (9.8%) | 47,592 (12.8%) |
| Depression/Anxiety/Bipolar | 110 (14.4%) | 26,102 (20.0%) | 41,258 (17.1%) | 67,470 (18.1%) |
| Medications within 12 months prior to the procedure | | | | |
| Opioid medication | 173 (22.7%) | 38,131 (29.2%) | 66,214 (27.4%) | 104,518 (28.0%) |
| NSAID medication | 134 (17.6%) | 46,162 (35.4%) | 67,425 (27.9%) | 113,721 (30.5%) |
| Tramadol medication | 31 (4.1%) | 14,702 (11.3%) | 16,693 (6.9%) | 31,426 (8.4%) |
| Neuropathic pain medication | 57 (7.5%) | 18,142 (13.9%) | 18,402 (7.6%) | 36,601 (9.8%) |
| Interventions in the 12 months prior to the procedure | | | | |
| Oral corticosteroids | 103 (13.5%) | 25,275 (19.4%) | 45,837 (19.0%) | 71,215 (19.1%) |
| Corticosteroid injection | 164 (21.5%) | 70,804 (54.3%) | 93,947 (38.9%) | 164,915 (44.2%) |
| Number of corticosteroid injections | | | | |
| Mean (SD) | 0.3 (0.8) | 1.1 (1.4) | 0.6 (1.1) | 0.8 (1.2) |
| Median (Q1, Q3) | 0.0 (0.0, 0.0) | 1.0 (0.0, 2.0) | 0.0 (0.0, 1.0) | 0.0 (0.0, 1.0) |
| Min, Max | 0.0, 9.0 | 0.0, 28.0 | 0.0, 71.0 | 0.0, 71.0 |
| Hyaluronic acid injection | 53 (6.9%) | 18,117 (13.9%) | 6,547 (2.7%) | 24,717 (6.6%) |
| Number of hyaluronic acid injections | | | | |
| Mean (SD) | 0.2 (0.7) | 0.3 (1.0) | 0.1 (0.4) | 0.2 (0.7) |
| Median (Q1, Q3) | 0.0 (0.0, 0.0) | 0.0 (0.0, 0.0) | 0.0 (0.0, 0.0) | 0.0 (0.0, 0.0) |
| Min, Max | 0.0, 8.0 | 0.0, 20.0 | 0.0, 12.0 | 0.0, 20.0 |
| Physical therapy | 282 (37.0%) | 45,155 (34.6%) | 65,577 (27.1%) | 111,014 (29.8%) |
| Number of PT visits | | | | |
| Mean (SD) | 4.1 (10.0) | 2.4 (6.4) | 2.3 (6.3) | 2.3 (6.3) |
| Median (Q1, Q3) | 0.0 (0.0, 4.0) | 0.0 (0.0, 1.0) | 0.0 (0.0, 1.0) | 0.0 (0.0, 1.0) |
| Min, Max | 0.0, 153.0 | 0.0, 150.0 | 0.0, 146.0 | 0.0, 153.0 |
| Health seeking behavior in the 12 months prior to the procedure | | | | |
| Outpatient visit | 733 (96.1%) | 129,703 (99.4%) | 239,821 (99.2%) | 370,257 (99.3%) |
| Number of outpatient visits | | | | |
| Mean (SD) | 15.9 (15.9) | 18.2 (14.2) | 15.2 (13.1) | 16.3 (13.6) |
| Median (Q1, Q3) | 11.0 (5.0, 22.0) | 15.0 (9.0, 23.0) | 12.0 (7.0, 20.0) | 13.0 (7.0, 21.0) |
| Min, Max | 0.0, 162.0 | 0.0, 360.0 | 0.0, 364.0 | 0.0, 364.0 |
| ER visit | 74 (9.7%) | 15,200 (11.7%) | 30,435 (12.6%) | 45,709 (12.3%) |
| Number of ER visits | | | | |
| Mean (SD) | 0.1 (0.4) | 0.2 (0.8) | 0.2 (0.7) | 0.2 (0.7) |
| Median (Q1, Q3) | 0.0 (0.0, 0.0) | 0.0 (0.0, 0.0) | 0.0 (0.0, 0.0) | 0.0 (0.0, 0.0) |
| Min, Max | 0.0, 4.0 | 0.0, 159.0 | 0.0, 66.0 | 0.0, 159.0 |
| Chiropractic visit | 142 (18.6%) | 14,446 (11.1%) | 29,532 (12.2%) | 44,120 (11.8%) |
| Number of chiropractic visits | | | | |

| Characteristic | **Orthobiologic procedures** N = 763 | **TKA/PKA** N = 130,466 | **Meniscectomy** N = 241,731 | **Overall** N = 372,960 |
|---|---|---|---|---|
| Mean (SD) | 1.8 (7.4) | 0.9 (4.0) | 1.0 (3.9) | 1.0 (3.9) |
| Median (Q1, Q3) | 0.0 (0.0, 0.0) | 0.0 (0.0, 0.0) | 0.0 (0.0, 0.0) | 0.0 (0.0, 0.0) |
| Min, Max | 0.0, 153.0 | 0.0, 118.0 | 0.0, 139.0 | 0.0, 153.0 |
| Knee x-ray | 316 (41.4%) | 115,439 (88.5%) | 205,706 (85.1%) | 321,461 (86.2%) |
| MRI of lower joint | 303 (39.7%) | 24,483 (18.8%) | 210,032 (86.9%) | 234,818 (63.0%) |
| Knee x-ray or MRI of lower joint | 429 (56.2%) | 117,137 (89.8%) | 226,884 (93.9%) | 344,450 (92.4%) |
| Other severity proxies in the 12 months prior to the procedure | | | | |
| Osteoarthritis outpatient visit for the treated knee | 382 (50.1%) | 126,801 (97.2%) | 97,251 (40.2%) | 224,434 (60.2%) |
| Number of osteoarthritis outpatient visits | | | | |
| Mean (SD) | 2.0 (3.6) | 5.0 (4.1) | 1.2 (2.4) | 2.5 (3.6) |
| Median (Q1, Q3) | 1.0 (0.0, 3.0) | 4.0 (3.0, 6.0) | 0.0 (0.0, 2.0) | 1.0 (0.0, 4.0) |
| Min, Max | 0.0, 47.0 | 0.0, 115.0 | 0.0, 111.0 | 0.0, 115.0 |
| Number of OA outpatient visits for the treated knee | | | | |
| Mean (SD) | 1.5 (2.4) | 4.4 (3.5) | 1.0 (2.0) | 2.2 (3.1) |
| Median (Q1, Q3) | 1.0 (0.0, 2.0) | 4.0 (2.0, 6.0) | 0.0 (0.0, 1.0) | 1.0 (0.0, 3.0) |
| Min, Max | 0.0, 16.0 | 0.0, 91.0 | 0.0, 111.0 | 0.0, 111.0 |
| Outpatient visit with an 'M' dx code | 637 (83.5%) | 128,937 (98.8%) | 232,829 (96.3%) | 362,403 (97.2%) |
| Number of outpatient visits with an 'M' dx code | | | | |
| Mean (SD) | 8.2 (10.7) | 9.1 (8.9) | 7.1 (8.1) | 7.8 (8.5) |
| Median (Q1, Q3) | 4.0 (1.0, 10.0) | 6.0 (4.0, 11.0) | 4.0 (3.0, 8.0) | 5.0 (3.0, 9.0) |
| Min, Max | 0.0, 68.0 | 0.0, 153.0 | 0.0, 178.0 | 0.0, 178.0 |

### Appendix R. Healthcare Resource Utilization Outcomes Sensitivity Analysis – 12 months

| Characteristic | Orthobiologic procedures N = 763 | TKA/PKA N = 130,466 | p-value[a] | Meniscectomy N = 241,731 | p-value[a] |
|---|---|---|---|---|---|
| Number of outpatient visits | | | < 0.001 | | 0.04 |
| Mean (SD) | 3.9 (4.8) | 7.5 (6.6) | | 3.7 (5.1) | |
| Median (Q1, Q3) | 2.0 (1.0, 5.0) | 6.0 (3.0, 10.0) | | 2.0 (0.0, 5.0) | |
| Min, Max | 0.0, 33.0 | 0.0, 199.0 | | 0.0, 114.0 | |
| Number of physical therapy visits | | | < 0.001 | | < 0.001 |
| Mean (SD) | 6.8 (12.0) | 16.2 (14.9) | | 6.6 (10.2) | |
| Median (Q1, Q3) | 0.0 (0.0, 9.0) | 13.0 (4.0, 24.0) | | 2.0 (0.0, 9.0) | |
| Min, Max | 0.0, 132.0 | 0.0, 166.0 | | 0.0, 149.0 | |
| Knee x-ray | 94 (12.3%) | 123,141 (94.4%) | < 0.001 | 55,305 (22.9%) | < 0.001 |
| MRI of lower joint | 84 (11.0%) | 4,653 (3.6%) | < 0.001 | 27,022 (11.2%) | 0.93 |
| CT of lower extremity | 12 (1.6%) | 4,959 (3.8%) | 0.002 | 2,606 (1.1%) | 0.25 |
| Subsequent same side TKA/PKA | 14 (1.8%) | 13,366 (10.2%) | < 0.001 | 7,112 (2.9%) | 0.09 |
| Subsequent same side Meniscectomy | <11 | 148 (0.1%) | -- | 13,663 (5.7%) | < 0.001 |
| Subsequent same side ACL/MCL procedure | <11 | 87 (0.1%) | -- | 902 (0.4%) | -- |
| Subsequent orthobiologic procedure - any knee | 88 (11.5%) | -- | -- | -- | -- |
| Subsequent orthobiologic procedure – same knee | 61 (8.0%) | -- | -- | -- | -- |
| Deep vein thrombosis inpatient care | <11 | 880 (0.7%) | 0.24 | 1,170 (0.5%) | -- |
| Hematoma inpatient care | <11 | 282 (0.2%) | -- | 110 (0.0%) | -- |
| Infection inpatient care | <11 | 2,357 (1.8%) | < 0.001 | 1,348 (0.6%) | -- |
| Pulmonary embolism inpatient care | <11 | 674 (0.5%) | -- | 713 (0.3%) | -- |
| Other post-surgical embolism inpatient care | <11 | 76 (0.1%) | -- | 68 (0.0%) | -- |
| DVT, PE, or OE inpatient care | <11 | 1,333 (1.0%) | 0.12 | 1,536 (0.6%) | -- |
| Opioid medication | 203 (26.6%) | 66,000 (50.6%) | < 0.001 | 53,973 (22.3%) | 0.005 |

[a]Compared to orthobiologic procedures

**Appendix S. Healthcare Resource Utilization Outcomes Sensitivity Analysis – 24 months**

| Characteristic | **Orthobiologic procedures** N = 512 | **TKA/PKA** N = 84,891 | p-value[a] | **Meniscectomy** N = 175,012 | p-value[a] |
|---|---|---|---|---|---|
| Number of outpatient visits | | | < 0.001 | | 0.18 |
| Mean (SD) | 6.7 (7.6) | 11.4 (10.1) | | 6.7 (8.7) | |
| Median (Q1, Q3) | 4.0 (1.0, 9.5) | 8.0 (5.0, 15.0) | | 4.0 (1.0, 9.0) | |
| Min, Max | 0.0, 46.0 | 0.0, 163.0 | | 0.0, 188.0 | |
| Number of physical therapy visits | | | < 0.001 | | 0.31 |
| Mean (SD) | 10.5 (21.1) | 18.7 (18.5) | | 8.7 (13.8) | |
| Median (Q1, Q3) | 2.0 (0.0, 14.0) | 15.0 (4.0, 27.0) | | 3.0 (0.0, 12.0) | |
| Min, Max | 0.0, 315.0 | 0.0, 282.0 | | 0.0, 266.0 | |
| Knee x-ray | 120 (23.4%) | 81,358 (95.8%) | < 0.001 | 58,395 (33.4%) | < 0.001 |
| MRI of lower joint | 96 (18.8%) | 5,269 (6.2%) | < 0.001 | 29,694 (17.0%) | 0.31 |
| CT of lower extremity | 12 (2.3%) | 4,374 (5.2%) | 0.006 | 3,262 (1.9%) | 0.52 |
| Subsequent same side TKA/PKA | 23 (4.5%) | 9,234 (10.9%) | < 0.001 | 8,824 (5.0%) | 0.64 |
| Subsequent same side Meniscectomy | 11 (2.1%) | 173 (0.2%) | < 0.001 | 11,657 (6.7%) | < 0.001 |
| Subsequent same side ACL/MCL procedure | <11 | 64 (0.1%) | -- | 802 (0.5%) | -- |
| Subsequent orthobiologic procedure - any knee | 70 (13.7%) | -- | -- | -- | -- |
| Subsequent orthobiologic procedure – same knee | 54 (10.5%) | -- | -- | -- | -- |
| Deep vein thrombosis inpatient care | <11 | 713 (0.8%) | -- | 1,112 (0.6%) | -- |
| Hematoma inpatient care | <11 | 229 (0.3%) | -- | 131 (0.1%) | -- |
| Infection inpatient care | <11 | 2,386 (2.8%) | 0.008 | 1,824 (1.0%) | 0.72 |
| Pulmonary embolism inpatient care | <11 | 618 (0.7%) | -- | 752 (0.4%) | -- |
| Other post-surgical embolism inpatient care | <11 | 64 (0.1%) | -- | 56 (0.0%) | -- |
| DVT, PE, or OE inpatient care | <11 | 1,134 (1.3%) | 0.1 | 1,528 (0.9%) | -- |
| Opioid medication | 172 (33.6%) | 44,810 (52.8%) | < 0.001 | 51,797 (29.6%) | 0.05 |

[a]Compared to orthobiologic procedures

**Appendix T. Healthcare Resource Utilization Outcomes Sensitivity Analysis – 36 months**

| Characteristic | Orthobiologic procedures N = 345 | TKA/PKA N = 55,015 | p-value[a] | Meniscectomy N = 125,649 | p-value[a] |
|---|---|---|---|---|---|
| Number of outpatient visits | | | < 0.001 | | 0.17 |
| Mean (SD) | 9.8 (11.1) | 14.9 (13.7) | | 9.6 (12.1) | |
| Median (Q1, Q3) | 6.0 (2.0, 14.0) | 11.0 (6.0, 19.0) | | 6.0 (2.0, 13.0) | |
| Min, Max | 0.0, 78.0 | 0.0, 187.0 | | 0.0, 292.0 | |
| Number of physical therapy visits | | | < 0.001 | | 0.76 |
| Mean (SD) | 14.0 (33.6) | 20.8 (21.5) | | 10.6 (17.1) | |
| Median (Q1, Q3) | 4.0 (0.0, 18.0) | 16.0 (5.0, 29.0) | | 4.0 (0.0, 14.0) | |
| Min, Max | 0.0, 532.0 | 0.0, 340.0 | | 0.0, 416.0 | |
| Knee x-ray | 119 (34.5%) | 53,006 (96.3%) | < 0.001 | 50,481 (40.2%) | 0.04 |
| MRI of lower joint | 91 (26.4%) | 4,715 (8.6%) | < 0.001 | 26,672 (21.2%) | 0.02 |
| CT of lower extremity | <11 | 3,309 (6.0%) | 0.01 | 3,139 (2.5%) | -- |
| Subsequent same side TKA/PKA | 23 (6.7%) | 5,925 (10.8%) | 0.02 | 8,022 (6.4%) | 0.92 |
| Subsequent same side Meniscectomy | <11 | 170 (0.3%) | -- | 8,951 (7.1%) | 0.003 |
| Subsequent same side ACL/MCL procedure | <11 | 41 (0.1%) | -- | 661 (0.5%) | -- |
| Subsequent orthobiologic procedure - any knee | 46 (13.3%) | -- | -- | -- | -- |
| Subsequent orthobiologic procedure – same knee | 34 (9.9%) | -- | -- | -- | -- |
| Deep vein thrombosis inpatient care | <11 | 588 (1.1%) | -- | 927 (0.7%) | -- |
| Hematoma inpatient care | <11 | 171 (0.3%) | -- | 119 (0.1%) | -- |
| Infection inpatient care | <11 | 2,011 (3.7%) | 0.04 | 1,826 (1.5%) | -- |
| Pulmonary embolism inpatient care | <11 | 526 (1.0%) | -- | 700 (0.6%) | -- |
| Other post-surgical embolism inpatient care | <11 | 48 (0.1%) | -- | 52 (0.0%) | -- |
| DVT, PE, or OE inpatient care | <11 | 930 (1.7%) | 0.16 | 1,335 (1.1%) | -- |
| Opioid medication | 139 (40.3%) | 30,110 (54.7%) | < 0.001 | 43,726 (34.8%) | 0.04 |

[a]Compared to orthobiologic procedures

### Appendix U. Healthcare Resource Utilization Outcomes Sensitivity Analysis – 48 months

| Characteristic | Orthobiologic procedures N = 235 | TKA/PKA N = 34,686 | p-value[a] | Meniscectomy N = 88,357 | p-value[a] |
|---|---|---|---|---|---|
| Number of outpatient visits | | | < 0.001 | | 0.01 |
| Mean (SD) | 13.4 (13.5) | 18.4 (17.3) | | 12.3 (15.3) | |
| Median (Q1, Q3) | 9.0 (4.0, 19.0) | 13.0 (7.0, 24.0) | | 7.0 (3.0, 16.0) | |
| Min, Max | 0.0, 72.0 | 0.0, 243.0 | | 0.0, 395.0 | |
| Number of physical therapy visits | | | < 0.001 | | 0.16 |
| Mean (SD) | 15.7 (21.2) | 22.6 (24.1) | | 12.5 (20.3) | |
| Median (Q1, Q3) | 7.0 (0.0, 25.0) | 17.0 (5.0, 31.0) | | 5.0 (0.0, 16.0) | |
| Min, Max | 0.0, 119.0 | 0.0, 411.0 | | 0.0, 628.0 | |
| Knee x-ray | 104 (44.3%) | 33,542 (96.7%) | < 0.001 | 40,275 (45.6%) | 0.73 |
| MRI of lower joint | 71 (30.2%) | 3,754 (10.8%) | < 0.001 | 21,842 (24.7%) | 0.06 |
| CT of lower extremity | 11 (4.7%) | 2,313 (6.7%) | 0.28 | 2,768 (3.1%) | 0.24 |
| Subsequent same side TKA/PKA | 20 (8.5%) | 3,586 (10.3%) | 0.42 | 6,591 (7.5%) | 0.63 |
| Subsequent same side Meniscectomy | <11 | 134 (0.4%) | -- | 6,408 (7.3%) | 0.03 |
| Subsequent same side ACL/MCL procedure | <11 | 29 (0.1%) | -- | 513 (0.6%) | -- |
| Subsequent orthobiologic procedure - any knee | 31 (13.2%) | -- | -- | -- | -- |
| Subsequent orthobiologic procedure – same knee | 21 (8.9%) | -- | -- | -- | -- |
| Deep vein thrombosis inpatient care | <11 | 449 (1.3%) | -- | 788 (0.9%) | -- |
| Hematoma inpatient care | <11 | 115 (0.3%) | -- | 114 (0.1%) | -- |
| Infection inpatient care | <11 | 1,554 (4.5%) | 0.03 | 1,666 (1.9%) | -- |
| Pulmonary embolism inpatient care | <11 | 377 (1.1%) | -- | 618 (0.7%) | -- |
| Other post-surgical embolism inpatient care | <11 | 32 (0.1%) | -- | 44 (0.0%) | -- |
| DVT, PE, or OE inpatient care | <11 | 699 (2.0%) | -- | 1,155 (1.3%) | -- |
| Opioid medication | 110 (46.8%) | 19,690 (56.8%) | 0.003 | 34,377 (38.9%) | 0.02 |

[a]Compared to orthobiologic procedures
